# Development of an Accurate and Rapid Antigen Assay for COVID-19 Diagnostics Using Saliva

**DOI:** 10.1101/2022.07.10.22277467

**Authors:** Camille Troup, Debnath Mukhopadhyay, Tania Chakrabarty, Anup Madan, Sri Satyanarayana, Shreefal Mehta, Su Dwarakanath

## Abstract

The global outbreak of COVID-19 highlighted the need for rapid and accurate diagnostic testing to control the spread of this highly contagious disease (1-5). Here, we describe the nCoVega COVID-19 antigen rapid test (∼ 15min) that can detect the presence of the SARS-COV-2 virus particles from saliva sample on a portable device. The portable reader instrument, the Vega-200, has a small footprint and is designed for use at point of care settings. The test detects the fluorescence signal using wide-field illumination from antigen-antibody complexes captured on a special filter matrix (6). Results of this clinical evaluation of 183 subjects demonstrates that the nCoVega COVID-19 test performs at par with qRT-PCR tests (7) (gold standard) for both symptomatic and asymptomatic patients, with a strong inverse correlation between RFU (relative fluorescence units) and Ct counts (from RT-PCR) maintaining detection accuracy even at very low viral loads. The test has an analytical performance of 15.3 TCID50/mL, and 100% specificity for COVID-19 as compared to other human respiratory viruses, including other human coronaviruses. The working principle of this assay and test system can be used for developing other rapid, inexpensive antigen assays and it can offer an end-to-end, point-of-care solution to meet the continuous demand in tackling existing and emerging infectious diseases across the globe.

## 1. Introduction

The nCoVega COVID-19 Antigen test, developed by Kaya 17, Inc., is an antigen-capture assay intended for the qualitative detection of spike protein antigen from SARS-CoV-2 in saliva from individuals, as viral antigen is generally detectable in saliva during the acute phase of infection. Positive results indicate the presence of viral antigens. This antigen test can be performed by trained non-laboratory or laboratory technicians for point of care use or for general use in clinical labs and hospital settings. The test works on a simple portable instrument called Vega-200 using a custom cartridge, assay reagents, and Vega software. The workflow uses a QR code reader and printer for tracking patient information.

The vast majority of COVID-19 antigen tests utilize lateral flow technology, which has inherent limitations in terms of dynamic range, sensitivity and specificity (8-10). The current rapid antigen tests are approved for nasal or NP swab. Other approaches include magnetic force assisted ELISA, microfluidic ELISA and digital chromatographic immunoassays, all of which require costly and complex custom manufacturing.

The nCoVega test utilizes selective antigen-antibody complex capture via filtration on an inexpensive and easy to manufacture disposable cartridge, and wide-field fluorescence imaging designed to give results in a few minutes with very high sensitivity and specificity. The reader is designed to run on USB power and is portable, light with no moving parts. Test format is designed so that 30-50 tests per hour can be processed on one reader. The workflow for our end-to-end solution including the test and the system is shown below in figure 1.

**Figure 1:**
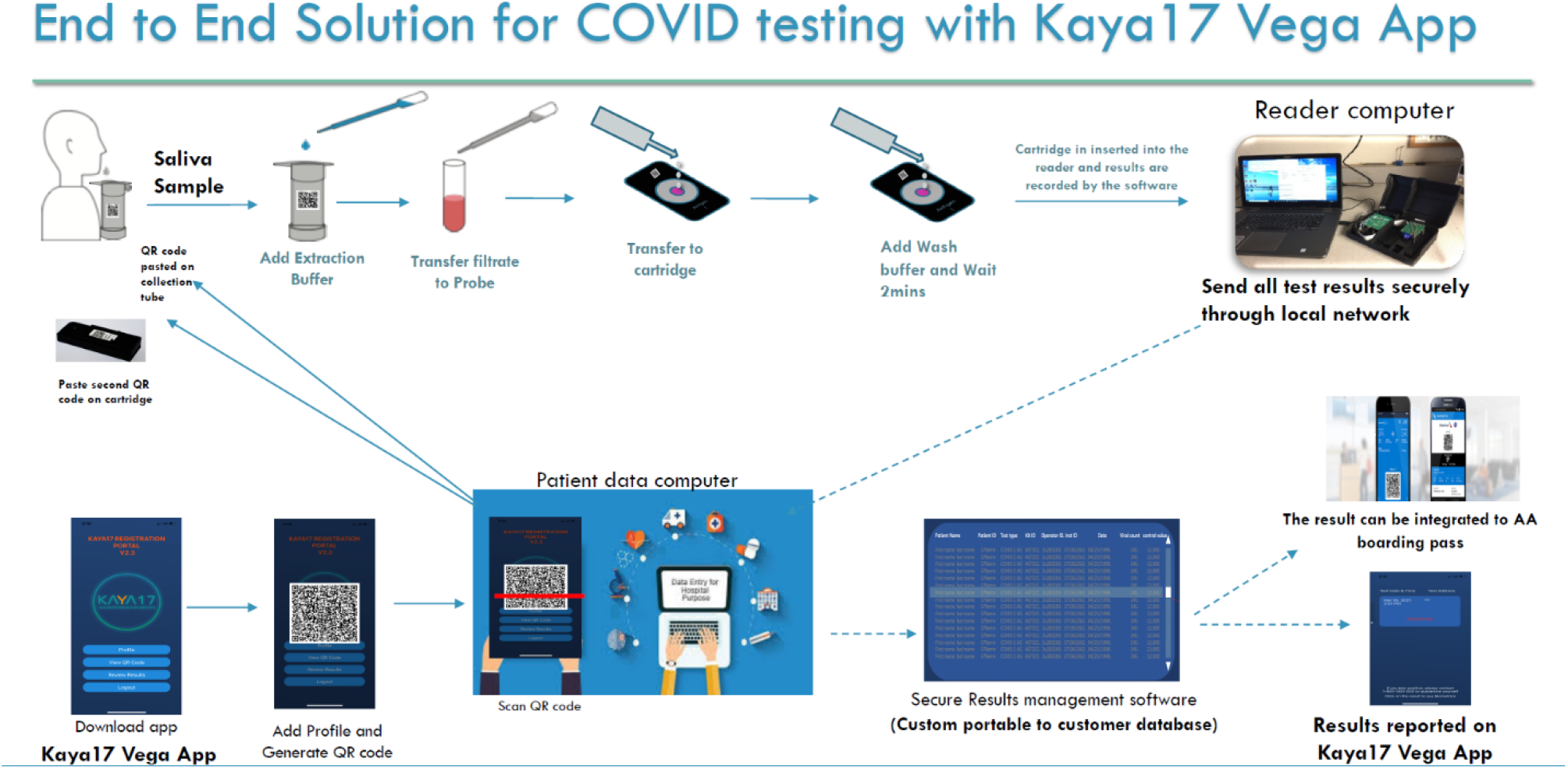
Schematic diagram showing the workflow for the nCoVega Test on the Vega-200 platform

The novel nCoVega COVID19 Antigen test is a direct dual probe antigen capture test using two fluorescently labeled primary antibodies (S1 and S2 spike protein recombinant IgG) that bind to SARS-CoV-2 spike protein on viral particles present in a saliva sample.

The developed test therefore can be easily deployed for large-scale population testing under community health settings, hospitals, and at the point of care with minimal operator training. This rapid and inexpensive test could help with COVID-19 diagnosis and controlling the spread of the global pandemic. The test can also be used to support decisions on infection control strategies such as detecting and isolating asymptomatic cases that could spread the virus unknowingly.

The study described here investigates the overall accuracy and sensitivity of the nCoVega antigen test in comparison with a high sensitivity RT-PCR method as the gold standard (7) for diagnosis of COVID-19.

## 2. Materials and Methods

Sample collection and processing: A saliva sample (approximately 200ul) is collected at the point of care testing by directly spitting into a vial that is placed in a syringe fitted with a filter. 1 mL of the proprietary sample processing reagent is added. A plunger is pushed to allow the treated saliva sample to pass through the filter and is collected in a second tube for antibody labeling. The filtrate is incubated with 30ul of fluorescently-labeled S1 and S2 antibodies for 10 minutes at room temperature. The mixture is then passed through a disposable vertical flow cartridge fitted with a proprietary capture matrix. The fluorescence signal from the captured antigen-antibody complex is recorded using a portable device equipped with wide-field illumination.

Antibodies and conjugation: The S1 and S2 antibodies used in our test are purchased from GeneTex, TX and conjugated with a Biotium Mix-n-Stain CF 405L fluorophore (Biotium Inc, Fremont, CA, USA). The fluorophore used in our test has a large Stokes Shift with an excitation peak at 395nm and emission peak around 545 nm. S1 and S2 antibodies are conjugated using the Mix-n-Stain protocol following the manufacturer’s protocol, which consists of direct labeling of the antibodies to the fluorophore via an NHS-ester covalent bond formation. The conjugated antibodies are then purified via spin column centrifugation and buffer exchanged into a PBS-based storage buffer. The CF dyes were carefully chosen for their superior brightness, photostability, large Stokes Shift, and resistance to hydrolysis offering superior performance over most dyes.

Instrument and data collection: The patient saliva sample labeled with fluorescent antibodies is placed on the cartridge inside a well, lined at the bottom with a capture matrix. An absorbent pad inside the cartridge holder to remove excess liquid and the unbound antibody. The bound antibody/antigen complex is selectively retained on the capture matrix via size exclusion mechanism. 500ul of wash reagent is added to the cartridge and allowed to wick through. Finally, 500ul of proprietary read reagent is added to the cartridge and the cartridge is immediately placed on the Vega-200 instrument for readout. The Vega-200 instrument records the fluorescence from the captured complex on the cartridge and reports a qualitative result (Positive or Negative). The system portable Vega-200 instrument is a fluorescence reader (U.S. Patent Application Serial Number: 16/690,589). The Vega analysis software, Patient ID software, an ID scanner with a QR code printer were all run on a standard PC. In addition, data reporting via a mobile device is also available.

### Data availability

Detailed data directly used to generate each figure or table of this study are available within the article, Supplementary Information and source data are also provided with this paper.

### Method used to determine LoD

Concentrations ranging from 1.6 × 105 TCID50/mL to 1.0 × 101 TCID50/mL Heat killed Zeptometrix SARS-CoV-2 (Cat. No. 0810587CFHI Lot No.324930) was spiked into artificial saliva matrix from Pickering laboratory (1700-0316).

Limit of Detection (LoD) for Kaya17 nCoVega test was determined by performing serial dilution (Table 1). A 10-fold serial dilution of was performed from 10,000, 1,000, 100, 10 and 0 of Heat Killed SARS CoV 2 from Zeptometrix in triplicate in Artificial saliva matrix from Pickering laboratory (1700-0316) (Figure 2a). From this, the LoD was estimated as 15.3 TCID50 per ml by Probit analysis (Figure 2b). Raw data is included in the supplementary material section (Table 1, supplementary materials section).

**Table 1:**
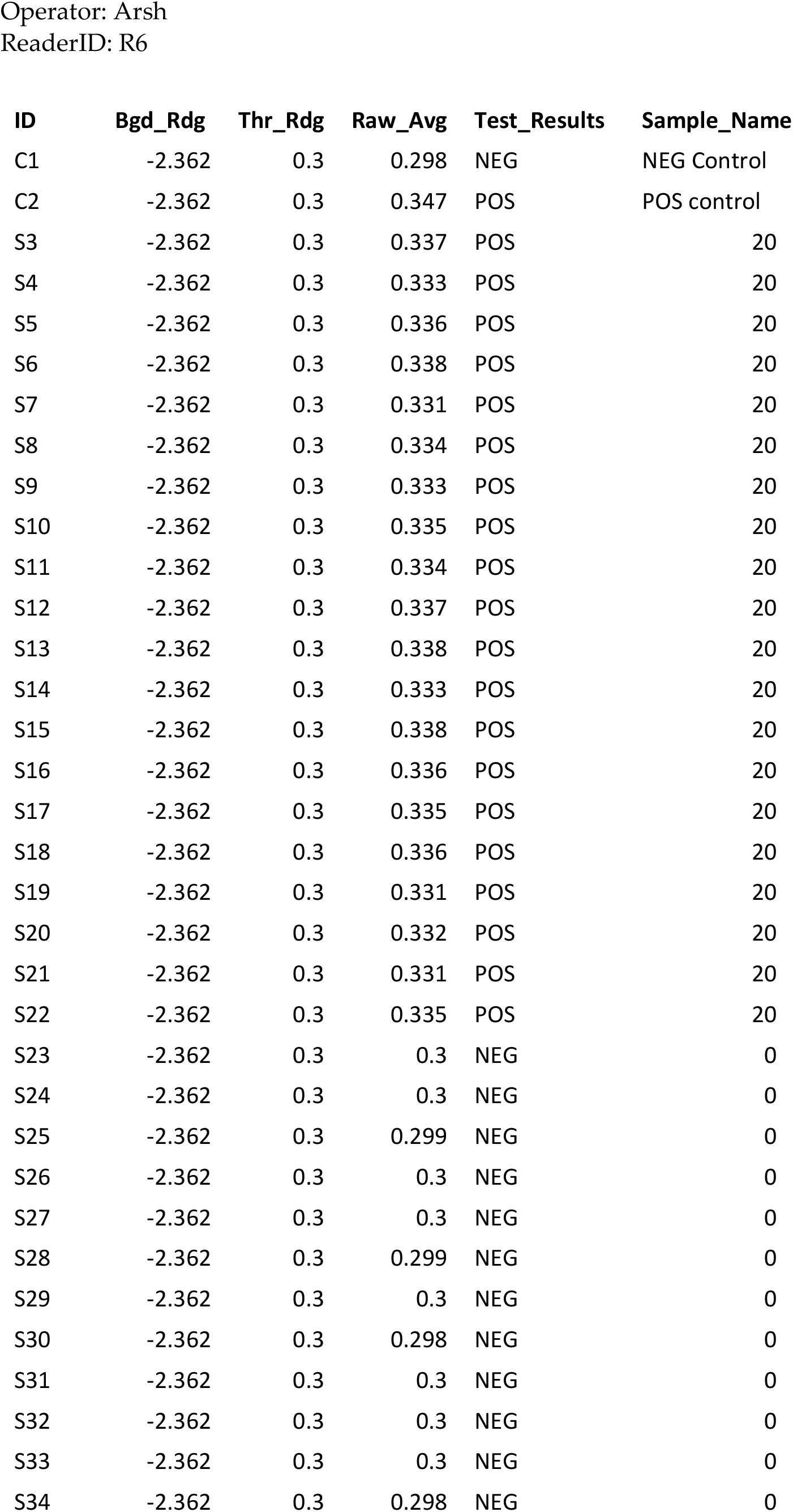
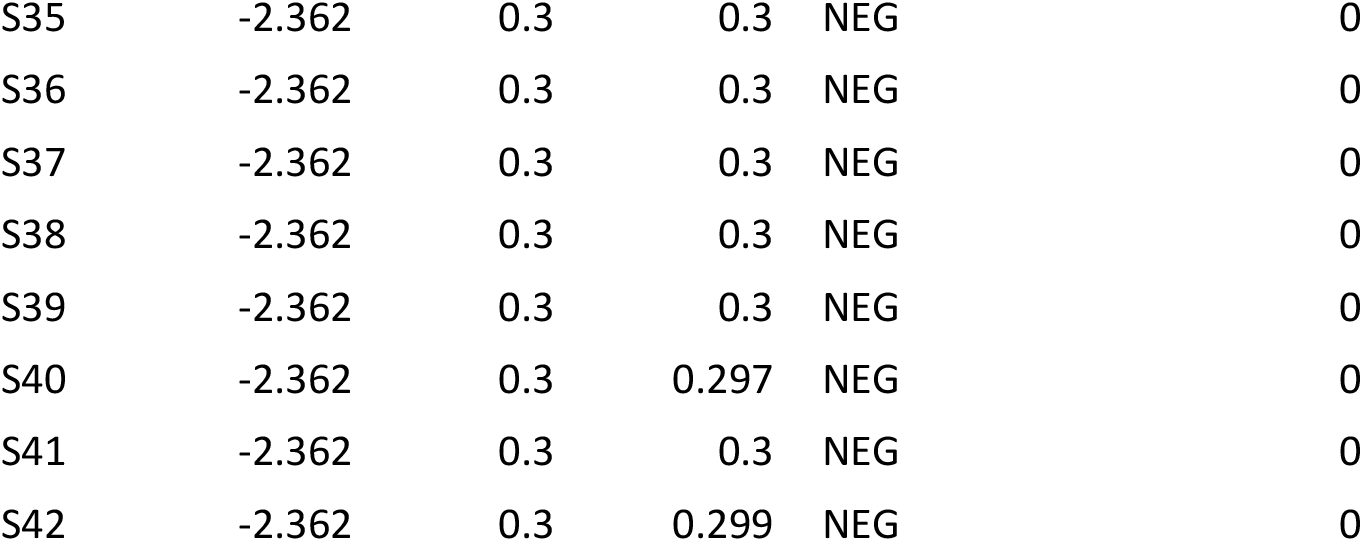
LOD LOB testing raw data.

| ID | Bgd_Rdg | Thr_Rdg | Raw_Avg | Test_Results | Sample_Name |
| --- | --- | --- | --- | --- | --- |
| C1 | -2.362 | 0.3 | 0.298 | NEG | NEG Control |
| C2 | -2.362 | 0.3 | 0.347 | POS | POS control |
| S3 | -2.362 | 0.3 | 0.337 | POS | 20 |
| S4 | -2.362 | 0.3 | 0.333 | POS | 20 |
| S5 | -2.362 | 0.3 | 0.336 | POS | 20 |
| S6 | -2.362 | 0.3 | 0.338 | POS | 20 |
| S7 | -2.362 | 0.3 | 0.331 | POS | 20 |
| S8 | -2.362 | 0.3 | 0.334 | POS | 20 |
| S9 | -2.362 | 0.3 | 0.333 | POS | 20 |
| S10 | -2.362 | 0.3 | 0.335 | POS | 20 |
| S11 | -2.362 | 0.3 | 0.334 | POS | 20 |
| S12 | -2.362 | 0.3 | 0.337 | POS | 20 |
| S13 | -2.362 | 0.3 | 0.338 | POS | 20 |
| S14 | -2.362 | 0.3 | 0.333 | POS | 20 |
| S15 | -2.362 | 0.3 | 0.338 | POS | 20 |
| S16 | -2.362 | 0.3 | 0.336 | POS | 20 |
| S17 | -2.362 | 0.3 | 0.335 | POS | 20 |
| S18 | -2.362 | 0.3 | 0.336 | POS | 20 |
| S19 | -2.362 | 0.3 | 0.331 | POS | 20 |
| S20 | -2.362 | 0.3 | 0.332 | POS | 20 |
| S21 | -2.362 | 0.3 | 0.331 | POS | 20 |
| S22 | -2.362 | 0.3 | 0.335 | POS | 20 |
| S23 | -2.362 | 0.3 | 0.3 | NEG | 0 |
| S24 | -2.362 | 0.3 | 0.3 | NEG | 0 |
| S25 | -2.362 | 0.3 | 0.299 | NEG | 0 |
| S26 | -2.362 | 0.3 | 0.3 | NEG | 0 |
| S27 | -2.362 | 0.3 | 0.3 | NEG | 0 |
| S28 | -2.362 | 0.3 | 0.299 | NEG | 0 |
| S29 | -2.362 | 0.3 | 0.3 | NEG | 0 |
| S30 | -2.362 | 0.3 | 0.298 | NEG | 0 |
| S31 | -2.362 | 0.3 | 0.3 | NEG | 0 |
| S32 | -2.362 | 0.3 | 0.3 | NEG | 0 |
| S33 | -2.362 | 0.3 | 0.3 | NEG | 0 |
| S34 | -2.362 | 0.3 | 0.298 | NEG | 0 |
| S35 | -2.362 | 0.3 | 0.3 | NEG | 0 |
| S36 | -2.362 | 0.3 | 0.3 | NEG | 0 |
| S37 | -2.362 | 0.3 | 0.3 | NEG | 0 |
| S38 | -2.362 | 0.3 | 0.3 | NEG | 0 |
| S39 | -2.362 | 0.3 | 0.3 | NEG | 0 |
| S40 | -2.362 | 0.3 | 0.297 | NEG | 0 |
| S41 | -2.362 | 0.3 | 0.3 | NEG | 0 |
| S42 | -2.362 | 0.3 | 0.299 | NEG | 0 |

**TABLE 1a:**
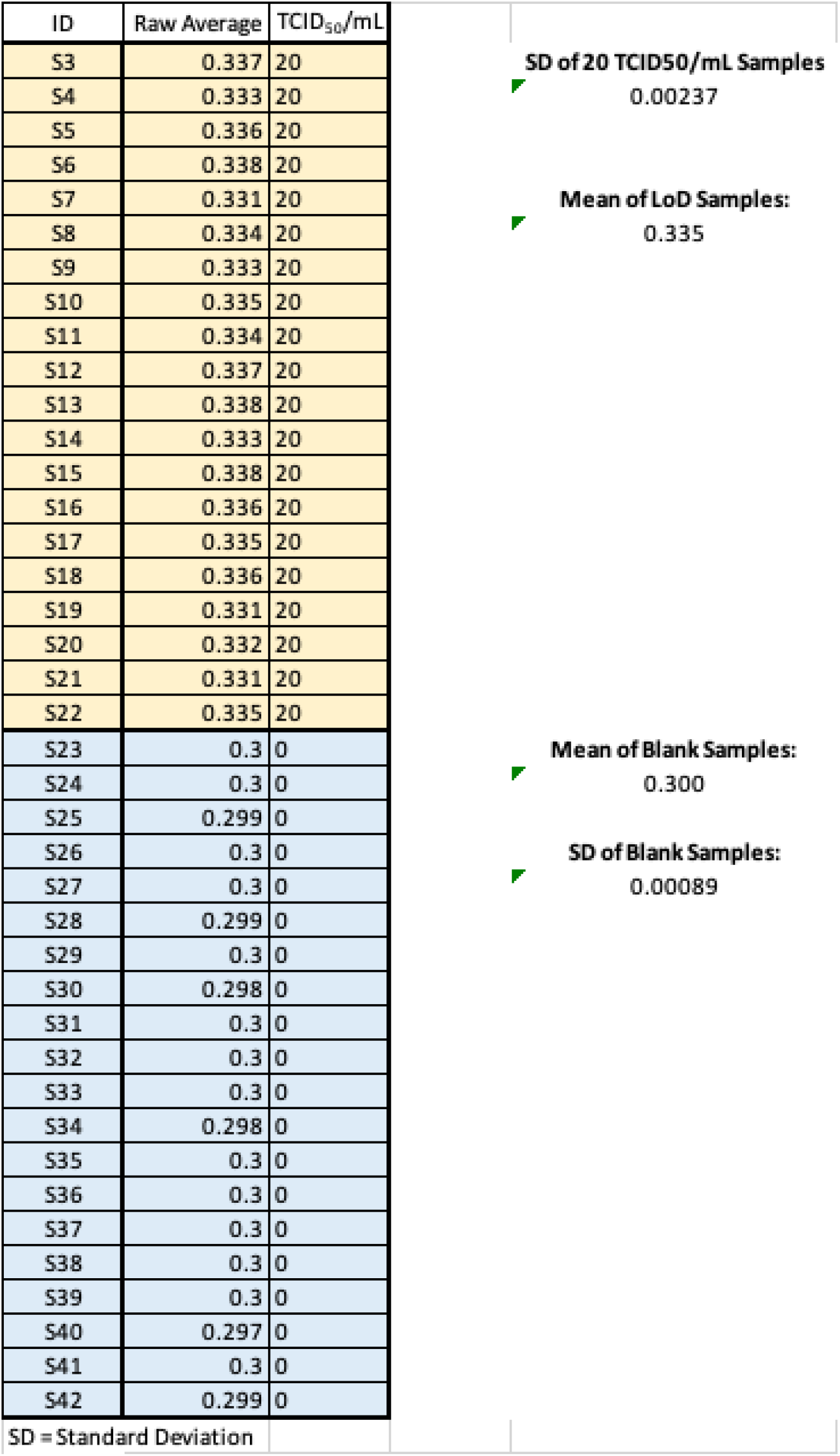
Results of 20 replicates for LoD and LoB confirmation.

| ID | Raw Average | TCID<sub>50</sub>/mL |  |
| --- | --- | --- | --- |
| S3 | 0.337 | 20 | **SD of 20 TCID50/mL Samples**<br>0.00237<br>**Mean of LoD Samples:**<br>0.335 |
| S4 | 0.333 | 20 |  |
| S5 | 0.336 | 20 |  |
| S6 | 0.338 | 20 |  |
| S7 | 0.331 | 20 |  |
| S8 | 0.334 | 20 |  |
| S9 | 0.333 | 20 |  |
| S10 | 0.335 | 20 |  |
| S11 | 0.334 | 20 |  |
| S12 | 0.337 | 20 |  |
| S13 | 0.338 | 20 |  |
| S14 | 0.333 | 20 |  |
| S15 | 0.338 | 20 |  |
| S16 | 0.336 | 20 |  |
| S17 | 0.335 | 20 |  |
| S18 | 0.336 | 20 |  |
| S19 | 0.331 | 20 |  |
| S20 | 0.332 | 20 |  |
| S21 | 0.331 | 20 |  |
| S22 | 0.335 | 20 |  |
| S23 | 0.3 | 0 | **Mean of Blank Samples:**<br>0.300<br>**SD of Blank Samples:**<br>0.00089 |
| S24 | 0.3 | 0 |  |
| S25 | 0.299 | 0 |  |
| S26 | 0.3 | 0 |  |
| S27 | 0.3 | 0 |  |
| S28 | 0.299 | 0 |  |
| S29 | 0.3 | 0 |  |
| S30 | 0.298 | 0 |  |
| S31 | 0.3 | 0 |  |
| S32 | 0.3 | 0 |  |
| S33 | 0.3 | 0 |  |
| S34 | 0.298 | 0 |  |
| S35 | 0.3 | 0 |  |
| S36 | 0.3 | 0 |  |
| S37 | 0.3 | 0 |  |
| S38 | 0.3 | 0 |  |
| S39 | 0.3 | 0 |  |
| S40 | 0.297 | 0 |  |
| S41 | 0.3 | 0 |  |
| S42 | 0.299 | 0 |  |
| SD = Standard Deviation |  |  |  |

**Table 2:**
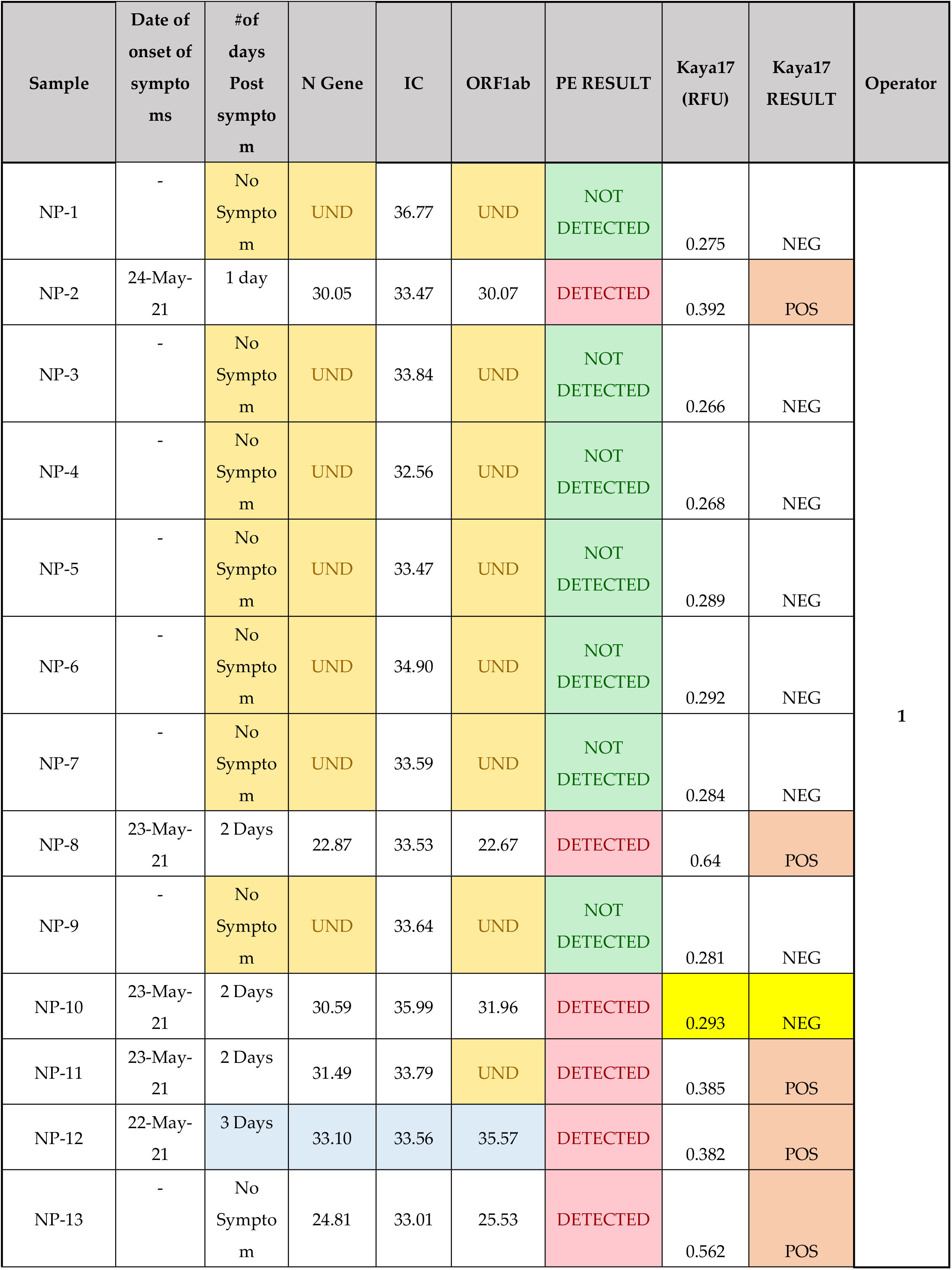
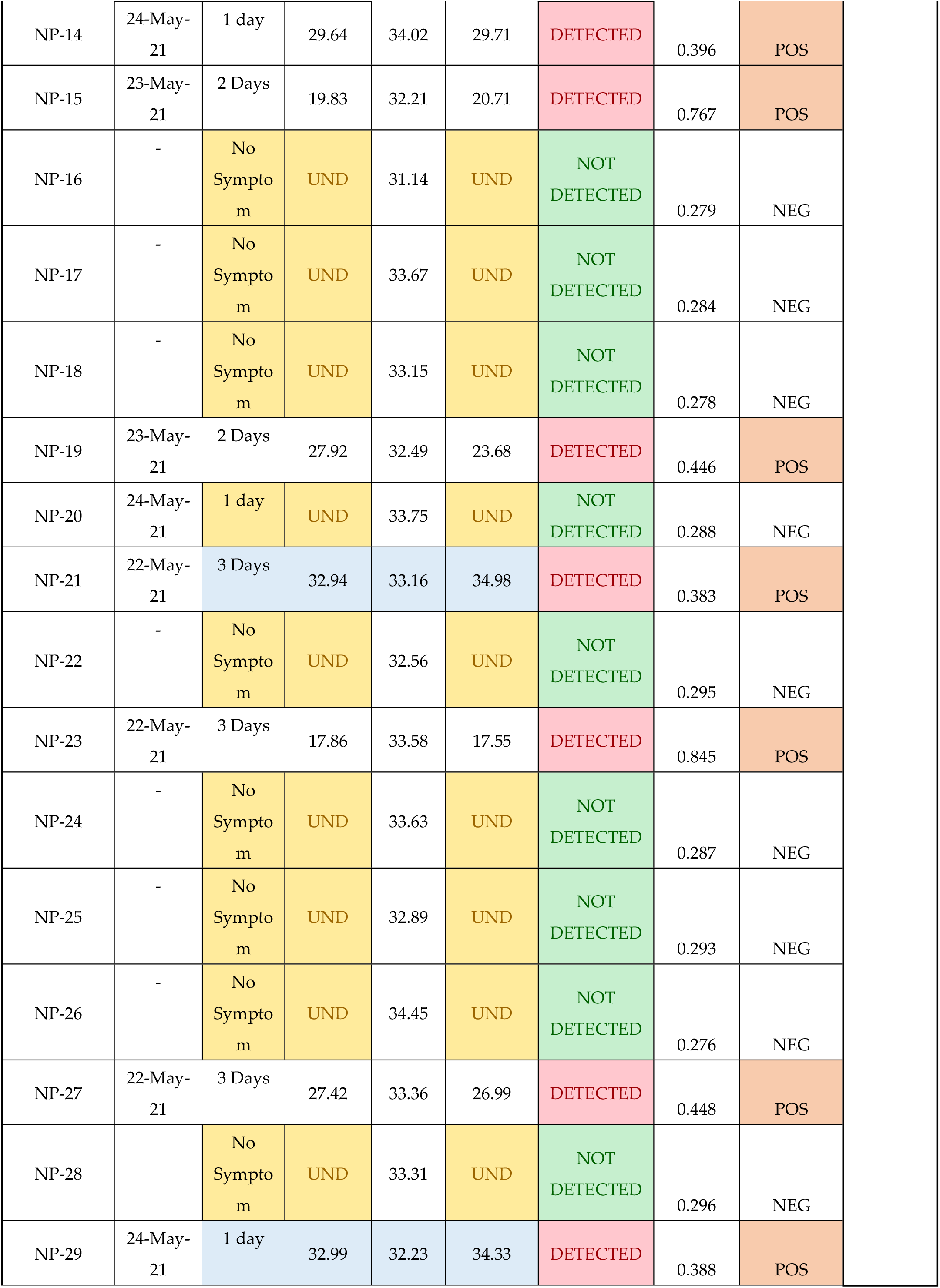
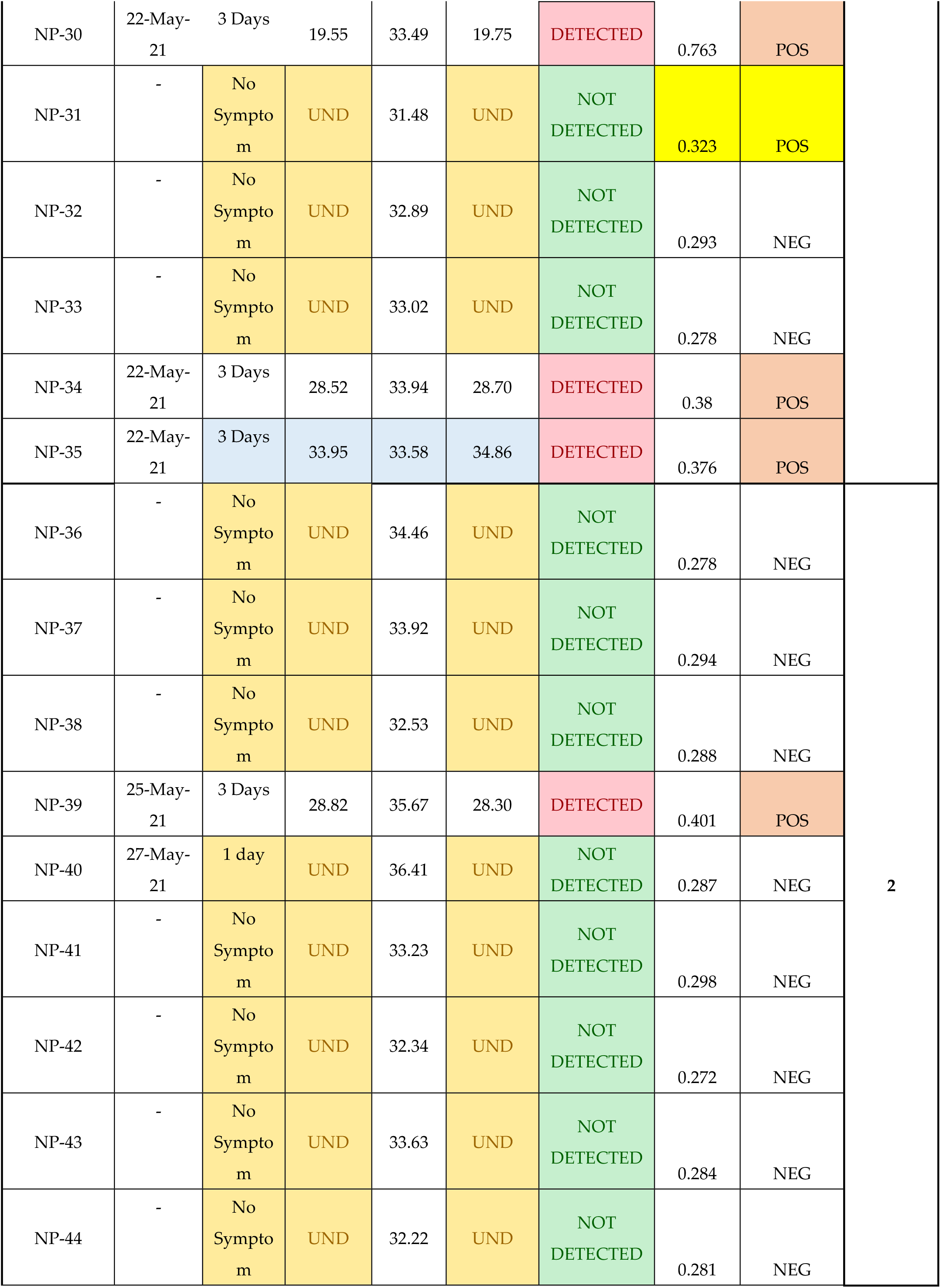
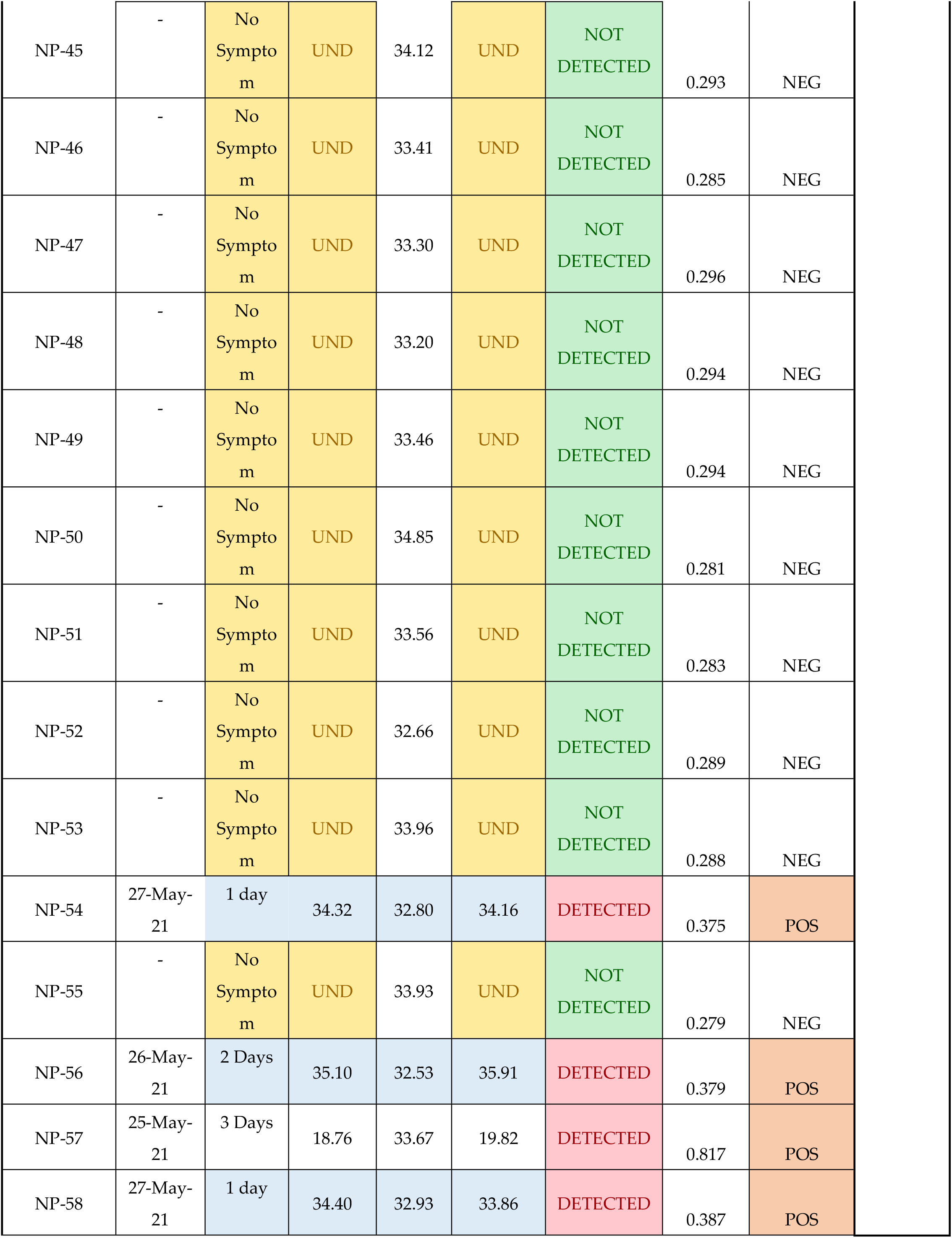
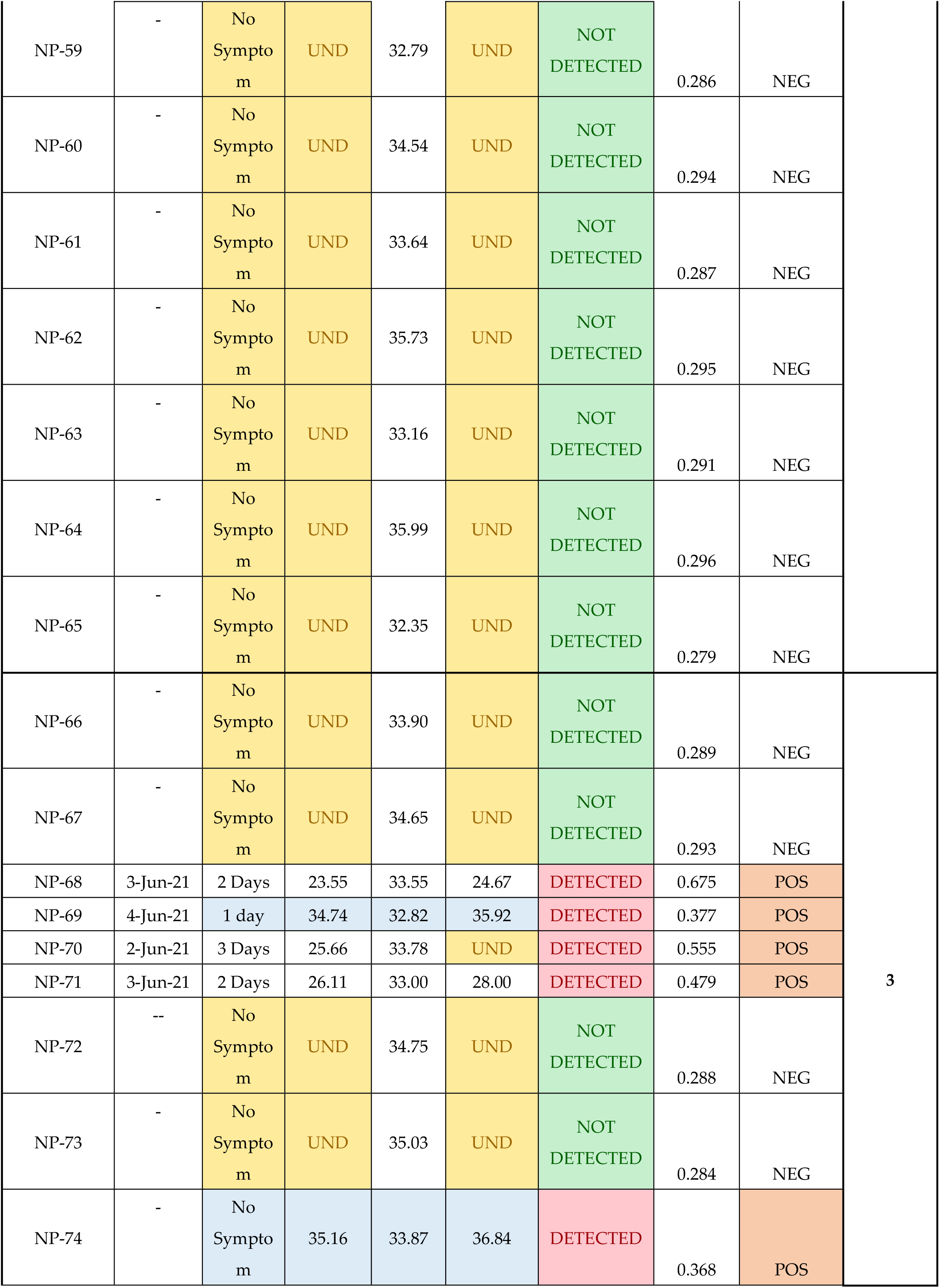
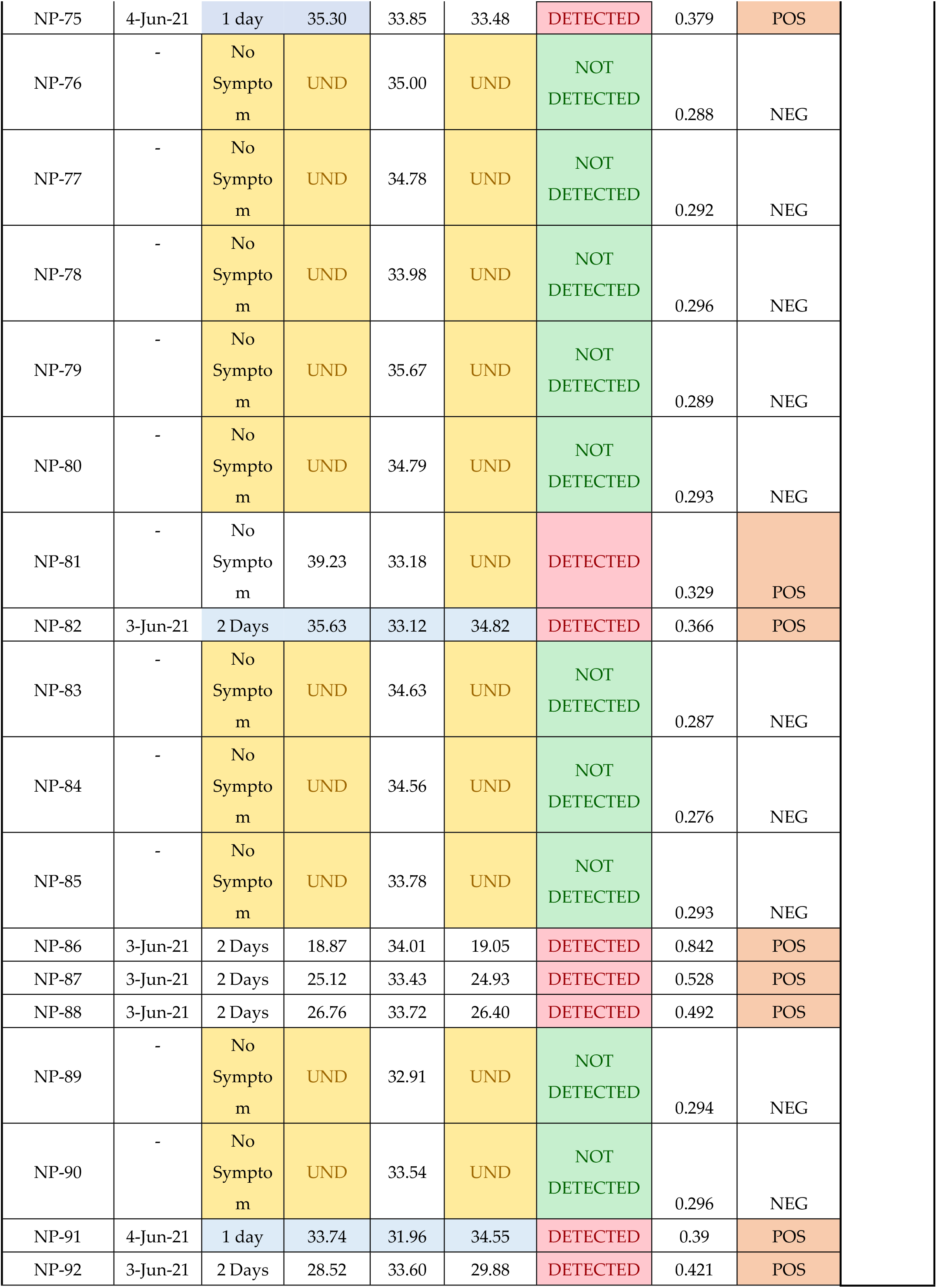
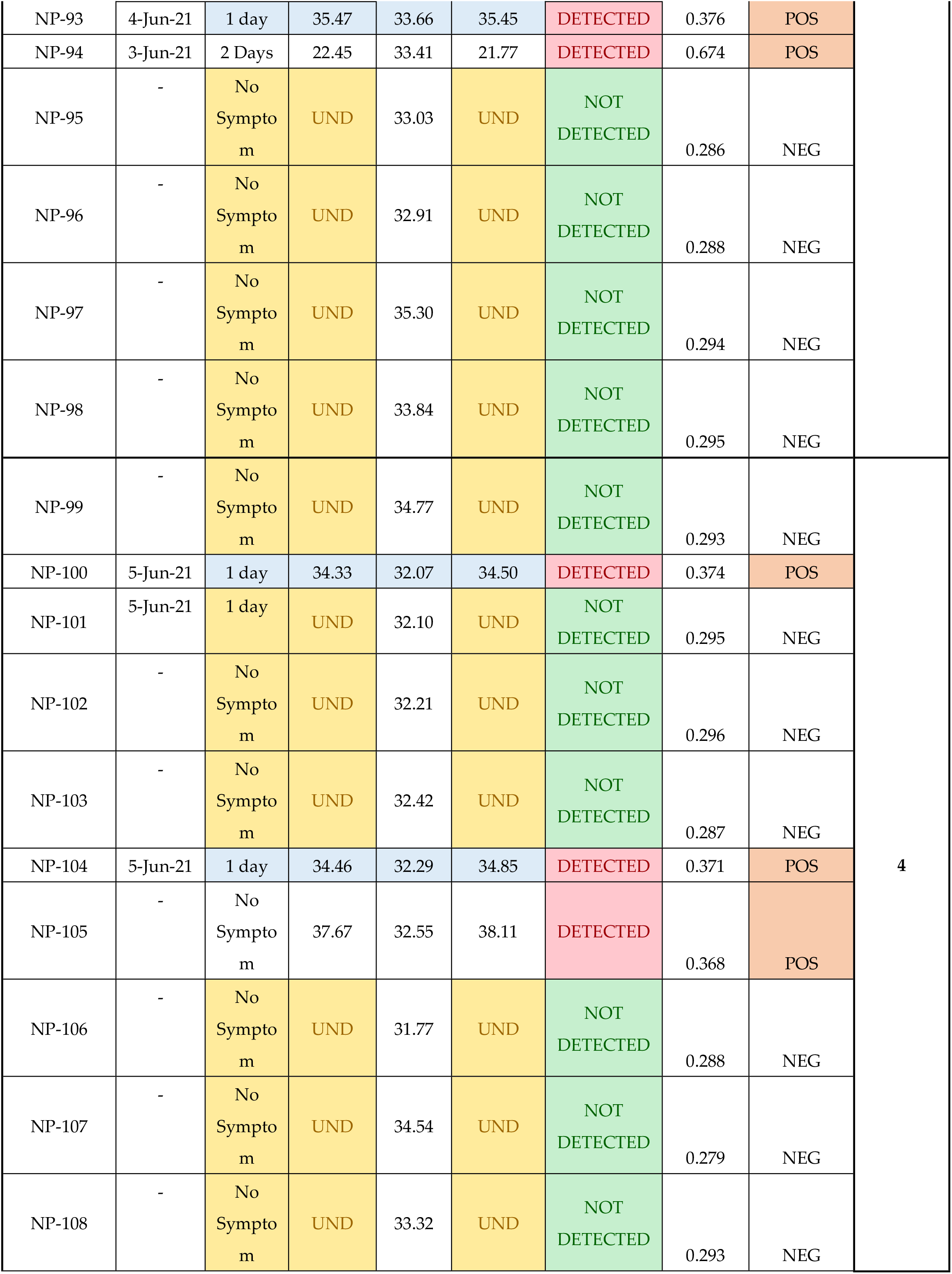
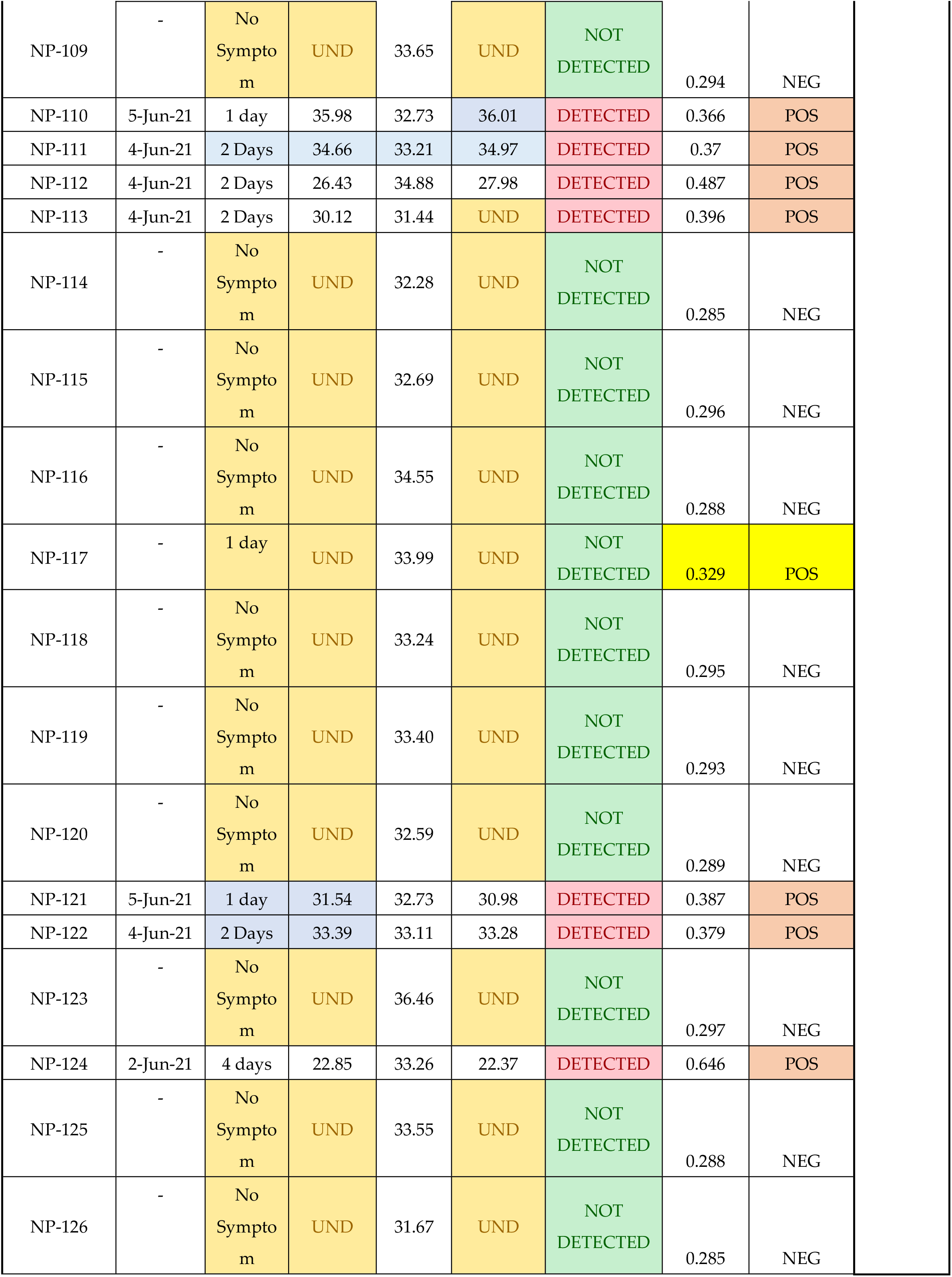
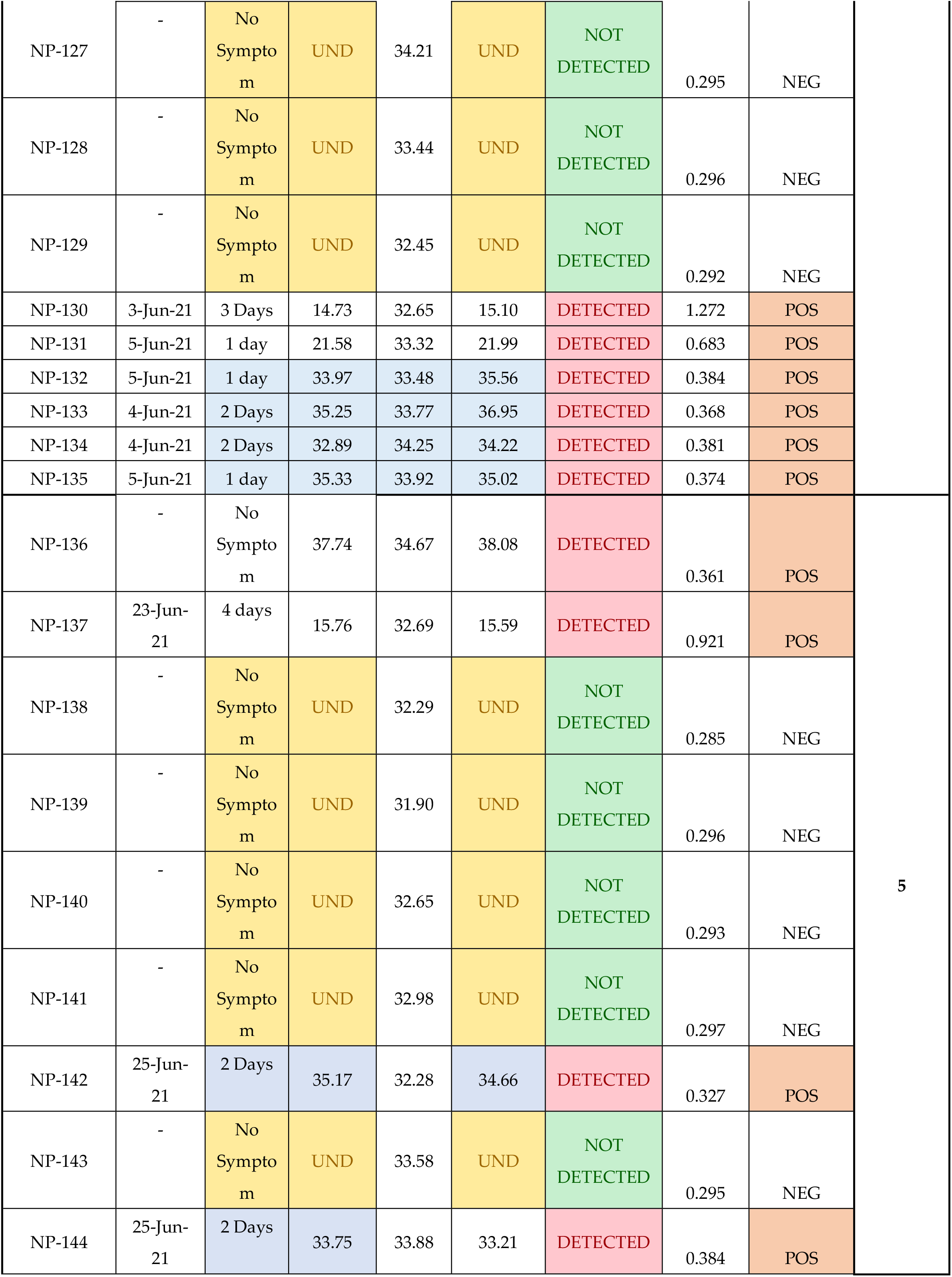
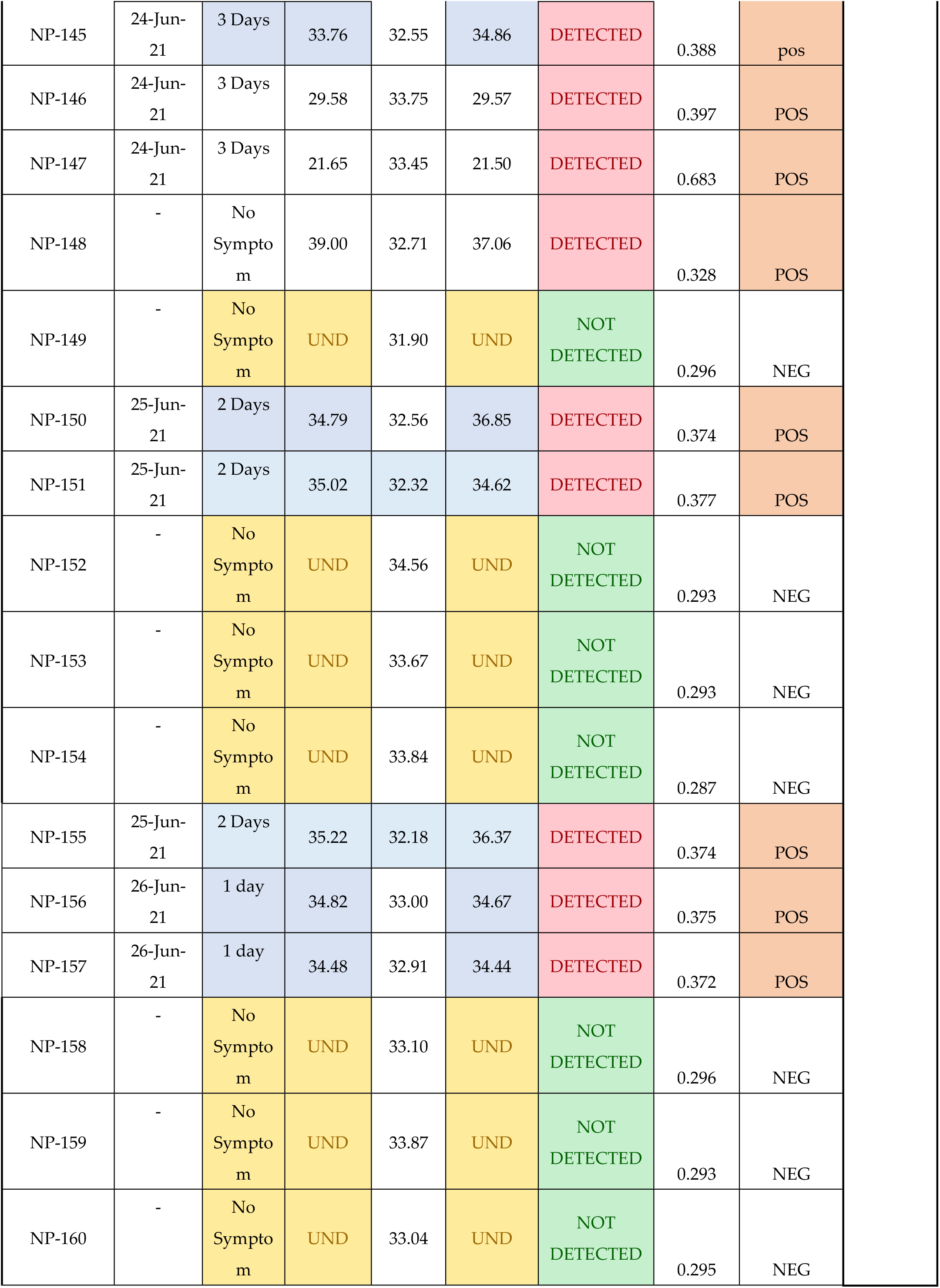
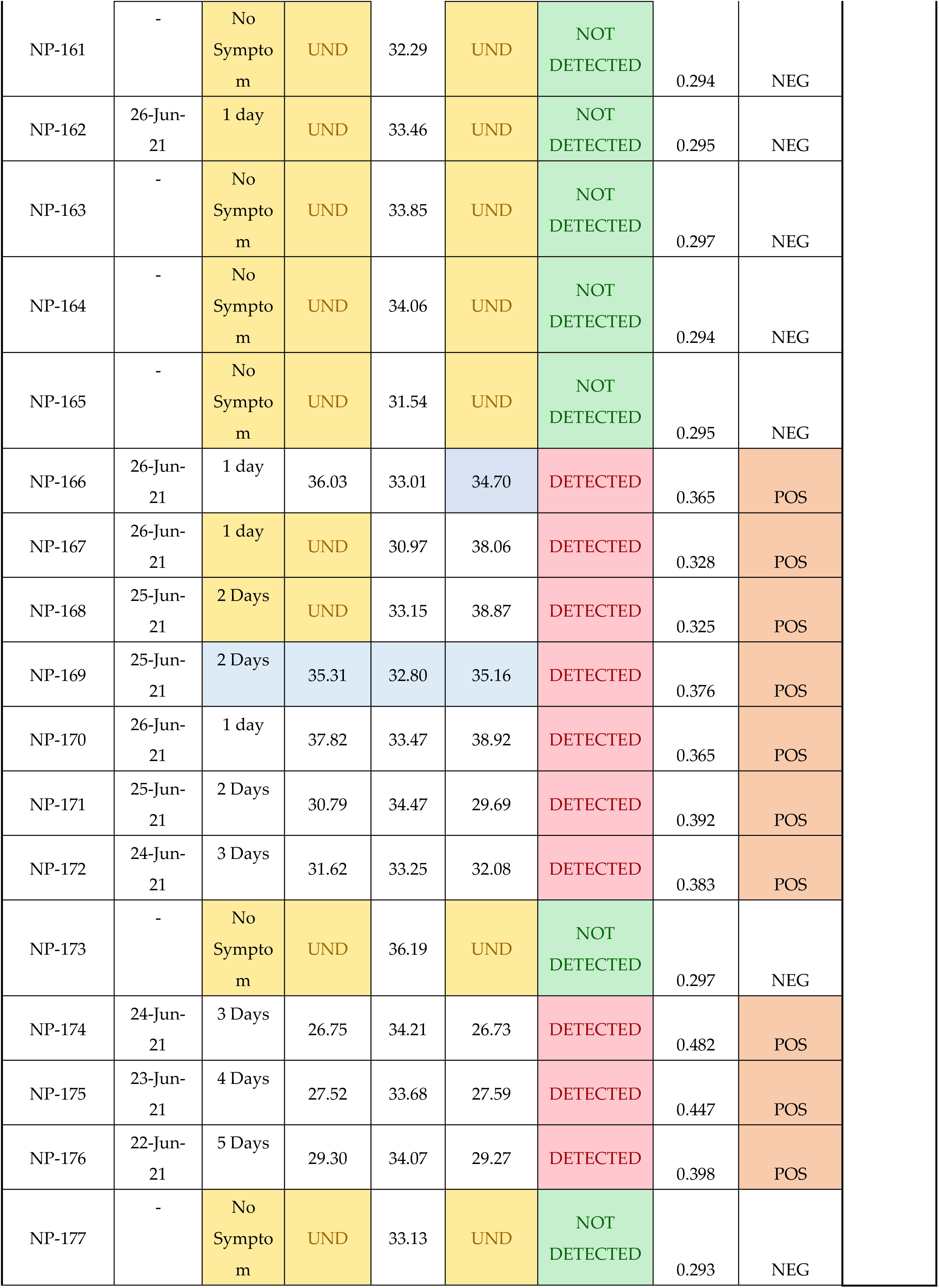
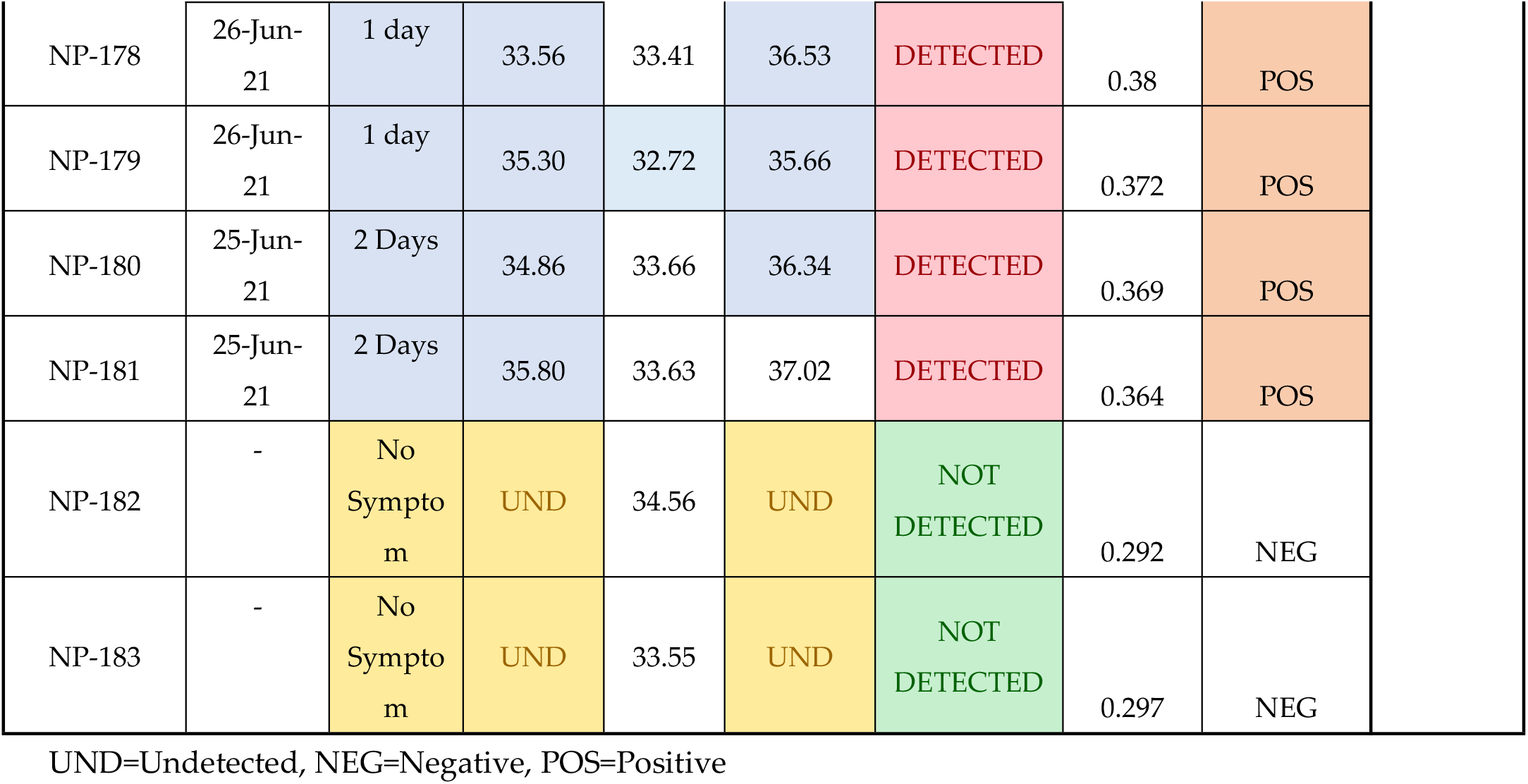
Clinical Test Results for PE Nucleic Acid Detection Kit and Kaya17 nCoVega Test.

**Figure 2.**
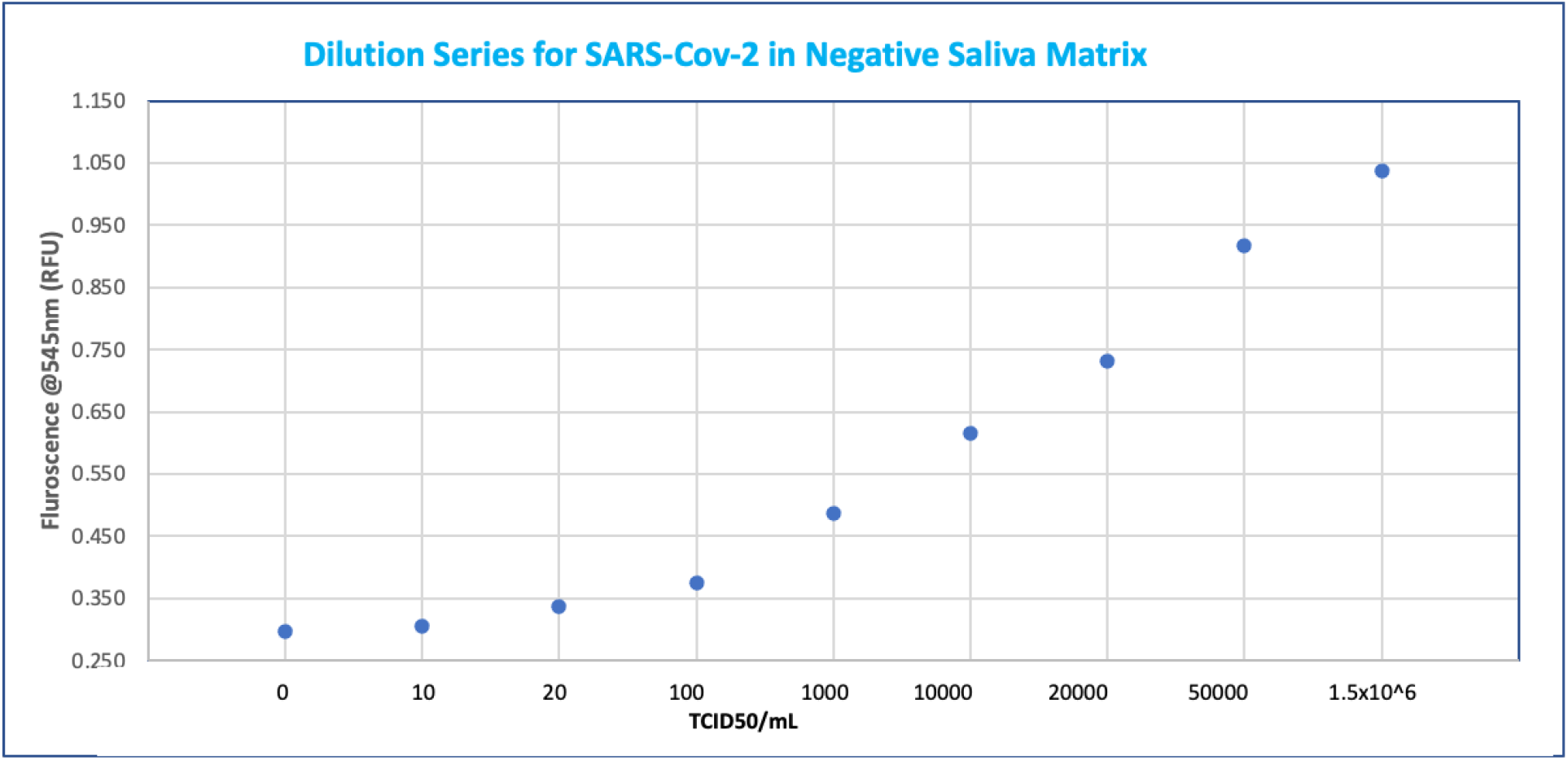
**(a):** LoD determination and dynamic range based on data from Table 1 (see supplementary section)

### Cross reactivity (analytical specificity)

Each organism was tested in the absence or presence of Heat Killed SARS-CoV-2 from Zeptometrix at 3 x LoD in Artificial saliva matrix from Pickering laboratory (1700-0316). The final concentration of the organisms is in the Table 3 below (the concentrations of 106 CFU/mL or higher for bacteria and 105 PFU/mL or higher for viruses is recommended). For some of the microorganisms, the stock concentration was lower than or equal to the recommended testing concentration. In such cases, these microorganisms were used at the stock concentration.

**Table 3:** Results of Cross-reactivity study.

| Microorganism | Source | Concentration | Cross-Reactivity (Yes/No) |
| --- | --- | --- | --- |
| Human coronavirus 229E | Zeptomatrix | $1 \times 10^5$ TCID<sub>50</sub>/mL | No (3/3 negative) |
| Human coronavirus OC43 | Zeptomatrix | $1 \times 10^5$ TCID<sub>50</sub>/mL | No (3/3 Negative) |
| Human coronavirus NL63 | Zeptomatrix | $1 \times 10^5$ TCID<sub>50</sub>/mL | No (3/3 Negative) |
| MERS coronavirus | Zeptomatrix | $1 \times 10^5$ TCID<sub>50</sub>/mL | No (3/3 Negative) |
| Adenovirus | Zeptomatrix | $1 \times 10^5$ TCID<sub>50</sub>/mL | No (3/3 Negative) |
| Human Metapneumovirus (hMPV) | Zeptomatrix | $1 \times 10^5$ TCID<sub>50</sub>/mL | No (3/3 Negative) |
| Parainfluenza virus Type 1 | Zeptomatrix | $1 \times 10^5$ TCID<sub>50</sub>/mL | No (3/3 Negative) |
| Parainfluenza virus Type 2 | Zeptomatrix | $1 \times 10^5$ TCID<sub>50</sub>/mL | No (3/3 Negative) |
| Parainfluenza virus Type 3 | Zeptomatrix | $1 \times 10^5$ TCID<sub>50</sub>/mL | No (3/3 Negative) |
| Parainfluenza virus Type 4a | Zeptomatrix | $1 \times 10^5$ TCID<sub>50</sub>/mL | No (3/3 Negative) |
| Influenza A H3N2 (Wisconsin/67/05) | BEI Resources | $5.4 \times 10^4$ TCID<sub>50</sub>/mL | No (3/3 Negative) |
| Influenza A H1N1 | Zeptomatrix | $1 \times 10^5$ TCID<sub>50</sub>/mL | No (3/3 Negative) |
| Influenza B (Florida/02/06) | Zeptomatrix | $1 \times 10^5$ TCID<sub>50</sub>/mL | No (3/3 Negative) |
| Enterovirus Type 68 | Zeptomatrix | $1 \times 10^5$ TCID<sub>50</sub>/mL | No (3/3 Negative) |
| Respiratory syncytial Virus Type B | Zeptomatrix | $1 \times 10^5$ TCID<sub>50</sub>/mL | No (3/3 Negative) |
| Rhinovirus | Zeptomatrix | $1 \times 10^5$ TCID<sub>50</sub>/mL | No (3/3 Negative) |
| SARS-CoV | BEI Resources | $1 \times 10^5$ PFU/mL | No (3/3 Negative) |
| Herpes Simplex Virus | Zeptomatrix | $1 \times 10^5$ TCID<sub>50</sub>/mL | No (3/3 Negative) |
| Epstein Barr Virus | Zeptomatrix | $1.03 \times 10^3$ cp/mL | No (3/3 Negative) |
| Haemophilus influenzae | Zeptomatrix | $1 \times 10^6$ CFU/mL | No (3/3 Negative) |
| Streptococcus pneumoniae | Zeptomatrix | $1 \times 10^6$ CFU/mL | No (3/3 Negative) |
| Streptococcus pyogenes | Zeptomatrix | $1 \times 10^6$ CFU/mL | No (3/3 Negative) |
| Candida albicans | Zeptomatrix | $1 \times 10^6$ CFU/mL | No (3/3 Negative) |
| Bordetella pertussis | Zeptomatrix | $1 \times 10^6$ CFU/mL | No (3/3 Negative) |
| Chlamydia pneumoniae | Zeptomatrix | $1 \times 10^6$ CFU/mL | No (3/3 Negative) |
| Legionella pneumophila | Zeptomatrix | $1 \times 10^6$ CFU/mL | No (3/3 Negative) |
| Mycobacterium tuberculosis | Zeptomatrix | $1 \times 10^6$ CFU/mL | No (3/3 Negative) |
| Pneumocystis jirovecii | Zeptomatrix | $1 \times 10^6$ CFU/mL | No (3/3 Negative) |
| Pseudomonas Aeruginosa | Zeptomatrix | $1 \times 10^6$ CFU/mL | No (3/3 Negative) |
| Staphylococcus Epidermidis | Zeptomatrix | $1 \times 10^6$ CFU/mL | No (3/3 Negative) |
| Streptococcus Salivarius | Zeptomatrix | $1 \times 10^6$ CFU/mL | No (3/3 Negative) |
| Mycoplasma pneumoniae | Zeptomatrix | $1 \times 10^6$ CFU/mL | No (3/3 Negative) |
| Pooled human Kaya17 nasal wash | Kaya17 | 14% v/v | No (3/3 Negative) |
| Elkenella corrodens | Zeptomatrix | $1 \times 10^6$ CFU/mL | No (3/3 Negative) |
| Streptococcus mutans | Zeptomatrix | $1 \times 10^6$ CFU/mL | No (3/3 Negative) |
| Staphylococcus aureus | Zeptomatrix | $1 \times 10^6$ CFU/mL | No (3/3 Negative) |
| Neisseria Meningitidis | Zeptomatrix | $1 \times 10^6$ CFU/mL | No (3/3 Negative) |
| Nocardia asteroides | Zeptomatrix | $1 \times 10^6$ CFU/mL | No (3/3 Negative) |
| Moraxella catarrhalis | Zeptomatrix | $1 \times 10^6$ CFU/mL | No (3/3 Negative) |
| Streptococcus mitis | Zeptomatrix | $1 \times 10^6$ CFU/mL | No (3/3 Negative) |

### Microbial Interference Studies

Each organism was tested in the absence or presence of Heat Killed SARS-CoV-2 from Zeptometrix at 3 x LoD in Artificial saliva matrix from Pickering laboratory (1700-0316). The final concentration of the organisms is in the Table 4 below (the concentrations of 106 CFU/mL or higher for bacteria and 10^5^ PFU/mL or higher for viruses is recommended). For some of the microorganisms, the stock concentration was lower than or equal to the recommended testing concentration. In such cases, these microorganisms were used at the stock concentration.

**Table 4:** Results of Microbial Interference studies

| Microorganism | Source | Concentration | Interference (Yes/No) |
| --- | --- | --- | --- |
| Human coronavirus 229E | Zeptomatrix | $1 \times 10^5$ TCID<sub>50</sub>/mL | No (3/3 positive) |
| Human coronavirus OC43 | Zeptomatrix | $1 \times 10^5$ TCID<sub>50</sub>/mL | No (3/3 positive) |
| Human coronavirus NL63 | Zeptomatrix | $1 \times 10^5$ TCID<sub>50</sub>/mL | No (3/3 positive) |
| MERS coronavirus | Zeptomatrix | $1 \times 10^5$ TCID<sub>50</sub>/mL | No (3/3 positive) |
| Adenovirus | Zeptomatrix | $1 \times 10^5$ TCID<sub>50</sub>/mL | No (3/3 positive) |
| Human Metapneumovirus (hMPV) | Zeptomatrix | $1 \times 10^5$ TCID<sub>50</sub>/mL | No (3/3 positive) |
| Parainfluenza virus Type 1 | Zeptomatrix | $1 \times 10^5$ TCID<sub>50</sub>/mL | No (3/3 positive) |
| Parainfluenza virus Type 2 | Zeptomatrix | $1 \times 10^5$ TCID<sub>50</sub>/mL | No (3/3 positive) |
| Parainfluenza virus Type 3 | Zeptomatrix | $1 \times 10^5$ TCID<sub>50</sub>/mL | No (3/3 positive) |
| Parainfluenza virus Type 4a | Zeptomatrix | $1 \times 10^5$ TCID<sub>50</sub>/mL | No (3/3 positive) |
| Influenza A H3N2 (Wisconsin/67/05) | BEI Resources | $5.4 \times 10^4$ TCID<sub>50</sub>/mL | No (3/3 positive) |
| Influenza A H1N1 | Zeptomatrix | $1 \times 10^5$ TCID<sub>50</sub>/mL | No (3/3 positive) |
| Influenza B (Florida/02/06) | Zeptomatrix | $1 \times 10^5$ TCID<sub>50</sub>/mL | No (3/3 positive) |
| Enterovirus Type 68 | Zeptomatrix | $1 \times 10^5$ TCID<sub>50</sub>/mL | No (3/3 positive) |
| Respiratory syncytial Virus Type B | Zeptomatrix | $1 \times 10^5$ TCID<sub>50</sub>/mL | No (3/3 positive) |
| Rhinovirus | Zeptomatrix | $1 \times 10^5$ TCID<sub>50</sub>/mL | No (3/3 positive) |
| SARS-CoV | BEI Resources | $1 \times 10^5$ PFU/mL | No (3/3 positive) |
| Herpes Simplex Virus | Zeptomatrix | $1 \times 10^5$ TCID<sub>50</sub>/mL | No (3/3 positive) |
| Epstein Barr Virus | Zeptomatrix | $1.03 \times 10^3$ cp/mL | No (3/3 positive) |
| Haemophilus influenzae | Zeptomatrix | $1 \times 10^6$ CFU/mL | No (3/3 positive) |
| Streptococcus pneumoniae | Zeptomatrix | $1 \times 10^6$ CFU/mL | No (3/3 positive) |
| *Streptococcus pyogenes* | Zeptomatrix | 1 x 10<sup>6</sup> CFU/mL | No (3/3 positive) |
| *Candida albicans* | Zeptomatrix | 1 x 10<sup>6</sup> CFU/mL | No (3/3 positive) |
| *Bordetella pertussis* | Zeptomatrix | 1 x 10<sup>6</sup> CFU/mL | No (3/3 positive) |
| *Chlamydia pneumoniae* | Zeptomatrix | 1 x 10<sup>6</sup> CFU/mL | No (3/3 positive) |
| *Legionella pneumophila* | Zeptomatrix | 1 x 10<sup>6</sup> CFU/mL | No (3/3 positive) |
| *Mycobacterium tuberculosis* | Zeptomatrix | 1 x 10<sup>6</sup> CFU/mL | No (3/3 positive) |
| *Pneumocystis jirovecii* | Zeptomatrix | 1 x 10<sup>6</sup> CFU/mL | No (3/3 positive) |
| *Pseudomonas Aeruginosa* | Zeptomatrix | 1 x 10<sup>6</sup> CFU/mL | No (3/3 positive) |
| *Staphylococcus Epidermidis* | Zeptomatrix | 1 x 10<sup>6</sup> CFU/mL | No (3/3 positive) |
| *Streptococcus Salivarius* | Zeptomatrix | 1 x 10<sup>6</sup> CFU/mL | No (3/3 positive) |
| *Mycoplasma pneumoniae* | Zeptomatrix | 1 x 10<sup>6</sup> CFU/mL | No (3/3 positive) |
| Pooled human Kaya17 nasal wash | Kaya17 | 14% v/v | No (3/3 positive) |
| *Elkenella corrodens* | Zeptomatrix | 1 x 10<sup>6</sup> CFU/mL | No (3/3 positive) |
| *Streptococcus mutans* | Zeptomatrix | 1 x 10<sup>6</sup> CFU/mL | No (3/3 positive) |
| *Staphylococcus aureus* | Zeptomatrix | 1 x 10<sup>6</sup> CFU/mL | No (3/3 positive) |
| *Neisseria Meningitidis* | Zeptomatrix | 1 x 10<sup>6</sup> CFU/mL | No (3/3 positive) |
| *Nocardia asteroides* | Zeptomatrix | 1 x 10<sup>6</sup> CFU/mL | No (3/3 positive) |
| *Moraxella catarrhalis* | Zeptomatrix | 1 x 10<sup>6</sup> CFU/mL | No (3/3 positive) |
| *Streptococcus mitis* | Zeptomatrix | 1 x 10<sup>6</sup> CFU/mL | No (3/3 positive) |

### Endogenous Interference Studies

Each substance was tested in triplicate in the absence or presence of Heat Killed SARS-CoV-2 from Zeptometrix at 3 x LOD in Artificial saliva matrix from Pickering laboratory (1700-0316). Substances for testing were selected based on the respiratory specimen’s guidance in http://www.accessdata.fda.gov/cdrh_docs/reviews/K112177.pdf.

### Hook Effect Testing

To determine if the Kaya17 nCoVega Test suffers from any high dose Hook effect, increasing concentrations of Heat Killed SARS-CoV-2 from Zeptometrix at 3 x LoD, were tested up to a concentration of 1.6 × 105 TCID50/mL. The starting material was spiked into a pooled negative saliva matrix obtained from healthy donors and confirmed negative for SARS-CoV-2. At each dilution, 500μL samples were added to Extraction buffer and tested using the nCoVega test.

### Clinical Evaluation

A prospective, single center, comparative study for validation of a rapid point-of-care test for diagnosis of SARS-COV-2 infection was conducted to evaluate the clinical use of the Kaya17 nCoVega COVID-19 Antigen test for use with saliva specimens collected neat.

The Kaya17 nCoVega COVID-19 Antigen test was clinically validated by Infinity BiologiX (IBX) using 183 paired saliva and swab samples (79 positives and 104 negatives) collected by St. Rose Hospital, Hayward, CA as per KAYA-PROTOPOCT-007 Prospective Clinical Study Protocol. The Western IRB approval number for the clinical trial was IRB protocol #20204097. Informed consents were collected from each subject along with basic medical history. The samples utilized in the study were analyzed using an RT PCR assay (Control), PerkinElmer New Coronavirus Nucleic Acid Detection Kit.

The primary objective of the study was to demonstrate efficacy of the Kaya17 nCoVega COVID-19 Antigen test in a non-laboratory setting by verifying the sensitivity and specificity of the Kaya17 nCoVega COVID-19 Antigen test in detecting SARS-COV-2 in positive saliva samples (yields a “positive” result) and not detecting SARS-COV-2 in negative saliva samples (yields a “negative” result) when performed by non-laboratory personnel. The secondary objective in the same protocol was to demonstrate that non-laboratory healthcare providers can perform the Kaya17 nCoVega COVID-19 Antigen test accurately in the intended use environment to Support Point of Care (POC) Use. (Point of Care study). In addition, the accuracy of detection was assessed in asymptomatic and symptomatic patients from day 1 of symptoms.

The data generated from the clinical study were statistically analyzed as per the following techniques

- Positive percent agreement (PPA),
- Negative percent agreement (NPA),
- Overall percent agreement (OPA),
- Associated 95% Wilson score confidence intervals
- Concordance analysis of the Kaya17 nCoVega Antigen test RFU vs Ct values of PCR

Subject status (negative/positive) was determined using a receiver operating characteristic (ROC) curve cutoff analysis. The evaluation metrics included positive percent agreement (PPA), negative percent agreement (NPA), overall percent agreement (OPA), and the absolute value of the sensitivity minus the specificity.

Before testing any of the samples, sample cartridges with positive and negative controls were inserted on the Vega 200 instrument that recorded fluorescence from these samples in RFU. This allowed the Vega analysis software to decide the cut-off value to use when deciding on positive and negative COVID-19 calls for sample testing. Details of the result interpretation are cited below in their respective sections. Raw data for the plots are also included in the supplementary information section.

All test controls were examined prior to interpretation of results. If the controls were not valid, the samples were not analyzed using the kit/instrument. Additional kits or instruments are required to be validated prior to running samples. The test result was determined by cutoff values based on fluorescence measurements on the Vega-200 instrument (excitation at 395 nm and emission at 545 nm). A “POSITIVE” result is called by the Vega software if the fluorescence measurement of the sample is above baseline fluorescence that has been determined by running the negative control multiple times. If the fluorescence value is at or below baseline, a “NEGATIVE” result is called by the Vega software. The results are stored in a CSV file on the Vega Reader computer. Further details are provided in the supplementary section.

## 3. Results

For laboratory validation of the nCoVega COVID19 Antigen test, a number of different studies were conducted such as analytical performance testing, LOD, cross-reactivity, inclusion and exclusion panel testing and hook effect testing. The details of each of those tests served to establish the sensitivity and specificity of the assay and the results are presented below.

### 3.1. Analytical Performance

#### 3.1.1. Limit of Detection (LoD) Determination

LoD studies were performed to determine the lowest detectable concentration of SARS-CoV-2 at which approximately 95% of all (true positive) replicates test positive.

From the ten-fold dilution series results in Table 1, the LoD can be determined as at least 10 TCID50/mL, since all samples with concentrations from 100000 TCID50/mL down to 10 TCID50/mL were above the 0 TCID50/mL. At 10 TCID50/mL, one of the replicates showed negative result (0.299 (S6)). Hence the LoD was set at 20 TCID50/mL. Probit analysis not possible due to 100% assessment of positivity at all positive concentration levels.

The ability to detect SARS-CoV-2 with Kaya17 nCoVega test at 20 TCID_50_ per ml was further confirmed by testing 20 replicates (Table 1a in supplementary section).

Limit of Blank (LoB) experiments were performed by running 20 samples at 0 TCID50 per ml LOB (Table 1a in supplementary section).

#### 3.1.2 Assay Cutoff Determination

Determining the assay cutoff was a key part of the data analysis. The assay cutoff determination for positives and negatives is determined using the Currie’s method (11) and CLSI EP17 guidance (12) for our qualitative purposes as follows.

From the guidance, the LoB’ is defined as:

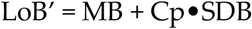

where M refers to the mean of “blank” samples and Cp is the multiplier and SDB is the Standard Deviation of the Blank Samples. As the data are very precise, in this case using a two-sided 5% significance level, Cp is defined as

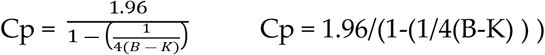

where B is the total number of blank replicates and K is the number of blank samples.

Therefore, from the data in the spreadsheet (see supplementary section), the LOB’ is calculated as:

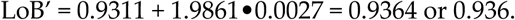

Next, per the guidance, the LOD’ can be calculated as:

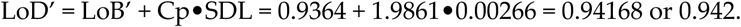

where Cp = 1.96/[1-(1/4•(Nreplicates – Nsamples))] = 1.96/[1-(1/4•(20-1))] = 1.9861.

and SDL = 0.00266 is the standard deviation of the positive low concentration samples with appropriate provision for degrees of freedom depending on the number of samples used to obtain the total replicates.

Therefore, if the LoB’ is taken as the cutoff (11), then we can establish a conservative equivocal zone between the LoB’ and the LoD’ as between 0.936 and 0.942 mean RR with the actual cutoff as 0.939 (as determined by ROC curve methodology in clinical evaluation (12)). Therefore, if a patient were to test in this equivocal zone, their result should be assigned as ‘Indeterminate’ and a retest is performed.

#### 3.1.3 Cross-reactivity (Analytical Specificity)

We performed cross-reactivity studies for Kaya17 nCoVega test using a panel of related pathogens, normal or pathogenic flora that are reasonably found in the clinical specimen. Also, the high prevalence disease agents likely to be encountered in the clinical specimen were tested for specificity of the test and the results showed no potential cross-reactivity with the Kaya17 nCoVega test with these agents, including various microorganisms and pathogens.

#### 3.1.4. Microbial Interference Studies

Microbial Interference for Kaya17 nCoVega test was evaluated by using a panel of related pathogens, normal or pathogenic flora that are reasonably found in the clinical specimen. Also, high prevalence disease agents were tested for specificity to demonstrate that false negatives do not occur when SARS-CoV-2 is present in a specimen with other microorganisms.

#### 3.1.5. Endogenous Interference Substances Studies

The following study was conducted to investigate whether potentially interfering substances, which may be found in the mouth and throat of symptomatic subjects (including over-the-counter medications), cross-react or interfere with the detection of SARS-CoV-2 using the Kaya17 nCoVega test. There was no interference with any of the tested materials in the study as described in Table 5.

**Table 5:** Results of Interference Studies

| Interfering Substance | Concentration | Interference (Yes/No) |
| --- | --- | --- |
| Clarispray | 50mcg | No |
|  |  | (3/3 Negative, 3/3 Positive) |
| Blood (human) | 5% | No |
|  |  | (3/3 Negative, 3/3 Positive) |
| Mucin | 5 mg/mL | No |
|  |  | (3/3 Negative, 3/3 Positive) |
| Fluticasone Propionate Flovont HFA | 110mcg | No |
|  |  | (3/3 Negative, 3/3 Positive) |
| Ventolin HFA Albuterol | 90mcg | No |
|  |  | (3/3 Negative, 3/3 Positive) |
| Budesonide and Formoterol Fumarate Dihydrate | 80mcg/4.5mcg | No |
|  |  | (3/3 Negative, 3/3 Positive) |
| Anefrin oxymetazoline HCl | 0.05% v/v | No |
|  |  | (3/3 Negative, 3/3 Positive) |
| Azelastine HCl | 137mcg | No |
|  |  | (3/3 Negative, 3/3 Positive) |
| Sore Throat Phenol Spray | 15% v/v | No |
|  |  | (3/3 Negative, 3/3 Positive) |
| Montelukast (Singular) | 0.5mg | No |
|  |  | (3/3 Negative, 3/3 Positive) |
| Loratadine (Claritin) | 0.15 mg/dL | No |
|  |  | (3/3 Negative, 3/3 Positive) |
| Human Genomic DNA | 10 ug.mL | No |
|  |  | (3/3 Negative, 3/3 Positive) |
| Nyquil (Acetaminophen, Doxylamine succinate, Dextromethorphan HBr) | 2 mg/mL | No |
|  |  | (3/3 Negative, 3/3 Positive) |
| Toothpaste (Colgate) | 0.5% v/v | No |
|  |  | (3/3 Negative, 3/3 Positive) |
| Crest/Listerine Mouthwash (Eucalyptol, menthol, Methyl Salicylate, Thymol) | 5% v/v | No<br>(3/3 Negative, 3/3 Positive) |
| Zinc | 10mg/mL | No<br>(3/3 Negative, 3/3 Positive) |
| Emergen-C | 10mg/mL | No<br>(3/3 Negative, 3/3 Positive) |
| Chloraseptic (Menthol/Benzocaine) | 1.5 mg/mL | No<br>(3/3 Negative, 3/3 Positive) |
| CVS Nasal Drops (Phenylephrine) | 15% v/v | No<br>(3/3 Negative, 3/3 Positive) |
| Afrin (Oxymetazoline) | 15% v/v | No<br>(3/3 Negative, 3/3 Positive) |
| CVS Nasal Spray (Cromolyn) | 15% v/v | No<br>(3/3 Negative, 3/3 Positive) |
| Zicam | 5% v/v | No<br>(3/3 Negative, 3/3 Positive) |
| Homeopathic (Alkalol) | 1:10 Dilution | No<br>(3/3 Negative, 3/3 Positive) |
| Sore Throat Phenol Spray | 15% v/v | No<br>(3/3 Negative, 3/3 Positive) |
| Robitussin | 2 mg/mL | No<br>(3/3 Negative, 3/3 Positive) |
| Alcohol (Ethanol) | 5% | No<br>(3/3 Negative, 3/3 Positive) |
| Nicotine | 0.03 mg/mL | No<br>(3/3 Negative, 3/3 Positive) |
| Vaseline (Petroleum Jelly) | 15% v/v | No<br>(3/3 Negative, 3/3 Positive) |
| Tamiflu (Oseltamivir Phosphate) | 5mg/mL | No<br>(3/3 Negative, 3/3 Positive) |
| Tobacco (Nicotine, Tar, Carbon Monoxide, Formaldehyde, Ammonia, Hydrogen Cyanide, Arsenic, and DDT) | 0.05mg/mL | No<br>(3/3 Negative, 3/3 Positive) |

### 3.2 High-dose Hook Effect

The Weibull statistical model was used for the data analysis (x axis=log10 TCID_50_ copies; y axis =Raw Average). The analysis showed that at the top end of the assay (1.6 × 10^5^ TCID_50_ copies) there is a plateau. This provides evidence that a Hook effect does not exist for this assay up to 1.6 × 10^5^ TCID_50_ copies. The model fits, as measured by the correlation and coefficient of determination, were each about 0.99 (closer to 1.00 is desired). Specific model information is shown in the figure 3.

**Figure 3.**
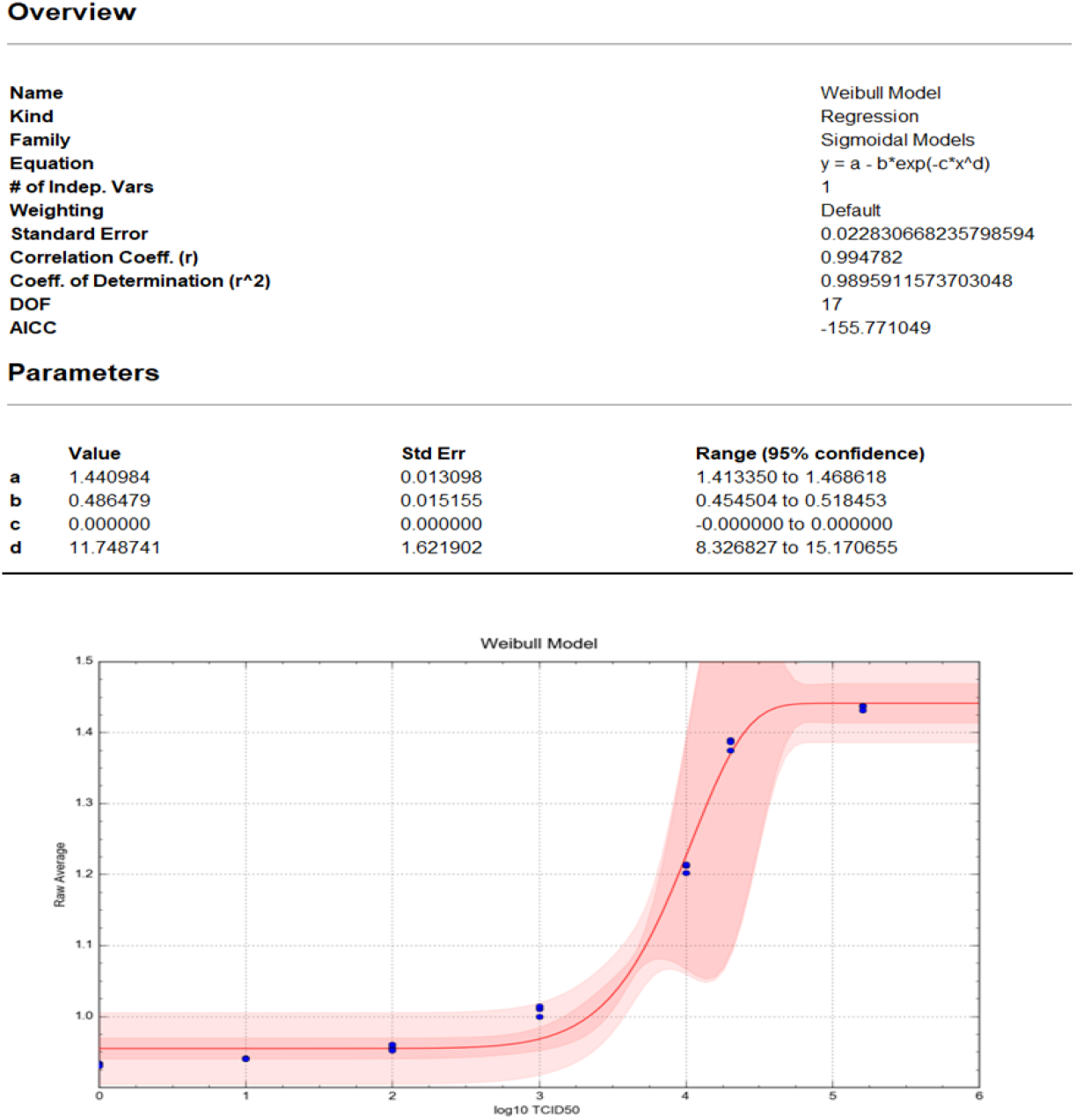
Weibull statistical analysis of Hook effect data

### 3.3 Clinical Evaluation

A total of n=183 samples were collected during the clinical evaluation. A total of n=183 test samples utilizing the subject device resulted in the same result (i.e., positive or negative) in comparison to the control, except two (n=2) test sample rendered a positive result when using the subject device and a negative result when using the control. One (n=1) test samples rendered a negative result when using the subject device and a positive result when using the control. After conducting statistical analysis, a 98.4% degree of accuracy is established based on the data set (Table 6).

**Table 6:** Accuracy in Overall Patient Population.

|  |  | PCR Reference Result |  |  | Agreement |  |  |
| --- | --- | --- | --- | --- | --- | --- | --- |
| Comparator Assay | Test Result | Positive | Negative | Total | Measure [a] | % (n/N) | 95% CI [b] |
| Kaya17 (Overall) | Positive | 78 | 2 | 80 | PPA | 98.7 (78/79) | (93.2, 99.8) |
|  | Negative | 1 | 102 | 103 | NPA | 98.1 (102/104) | (93.3, 99.5) |
|  | Total | 79 | 104 | 183 | OPA | 98.4 (180/183) | (95.3, 99.4) |
| [a] PPA=Positive Percent Agreement, NPA=Negative Percent Agreement, OPA=Overall Percent Agreement.<br>[b] Wilson Score Confidence Interval. |  |  |  |  |  |  |  |

#### Accuracy in Overall Patient Population

The test results include 79 True Positives (RT-PCR Positives) and 104 True negatives (RT-PCR Negatives) (data in Table 2 in supplemental materials). When we analyzed the performance of the Kaya17 assay in overall population, the PPA value is calculated to be 98.7% and at 95% Confidence Interval, values are found to be at (93.2% Lower bound, 99.8% Upper bound).

#### Accuracy in Symptomatic Patient Population

In the Symptomatic population (Table 7), there are 73 RT-PCR positives (True Positives) and 45 RT-PCR negatives (True Negatives). When we analyzed the performance of the Kaya17 assay in symptomatic population, the PPA value is calculated to be 98.6% and at *95% Confidence Interval*, values are found to be at (92.60% Lower bound, 99.8% Upper bound).

**Table 7:** Accuracy in Symptomatic Patient Population.

|  |  | PCR Reference Result |  |  | Agreement |  |  |
| --- | --- | --- | --- | --- | --- | --- | --- |
| Comparator Assay | Test Result | Positive | Negative | Total | Measure [a] | % (n/N) | 95% CI [b] |
| Kaya17 (Symptomatic) | Positive | 72 | 1 | 73 | PPA | 98.6 (72/73) | (92.6, 99.8) |
|  | Negative | 1 | 44 | 45 | NPA | 97.8 (44/45) | (88.4, 99.6) |
|  | Total | 73 | 45 | 118 | OPA | 98.3 (116/118) | (94.0, 99.5) |
| [a] PPA=Positive Percent Agreement, NPA=Negative Percent Agreement, OPA=Overall Percent Agreement.<br>[b] Wilson Score Confidence Interval. |  |  |  |  |  |  |  |

As seen in the raw data in Table 2 in the supplementary section, the nCovega test was able to detect the SARS-COV-2 virus as early as day 1 from the onset of smptoms.

#### Accuracy in Asymptomatic Patient Population

In the Asymptomatic population (Table 8), there are 6 RT-PCR positives (True Positives) and 59 RT-PCR negatives (True Negatives). When we analyzed the performance of the Kaya17 assay in asymptomatic population, the PPA value is calculated to be 100% and at *95% Confidence Interval*, values are found to be at (61% Lower bound, 100% Upper bound).

**Table 8:** Accuracy in Asymptomatic Patient Population.

| Comparator Assay | Test Result | PCR Reference Result |  |  | Agreement |  |  |
| --- | --- | --- | --- | --- | --- | --- | --- |
|  |  | Positive | Negative | Total | Measure [a] | % (n/N) | 95% CI [b] |
| Kaya17 (Asymptomatic) | Positive | 6 | 1 | 7 | PPA | 100.0<br>(6/6) | (61.0, 100.0) |
|  | Negative | 0 | 58 | 58 | NPA | 98.3<br>(58/59) | (91.0, 99.7) |
|  | Total | 6 | 59 | 65 | OPA | 98.5<br>(64/65) | (91.8, 99.7) |
| [a] PPA=Positive Percent Agreement, NPA=Negative Percent Agreement, OPA=Overall Percent Agreement.<br>[b] Wilson Score Confidence Interval. |  |  |  |  |  |  |  |

The operator specific accuracy is measured to support the POC Study to demonstrate that the Kaya17 nCoVega COVID-19 Antigen test can be performed by the untrained operator following the Quick Reference Guide without any training. Each untrained operator has achieved the 95% accuracy. Please refer to the Table 9.

**Table 9:**
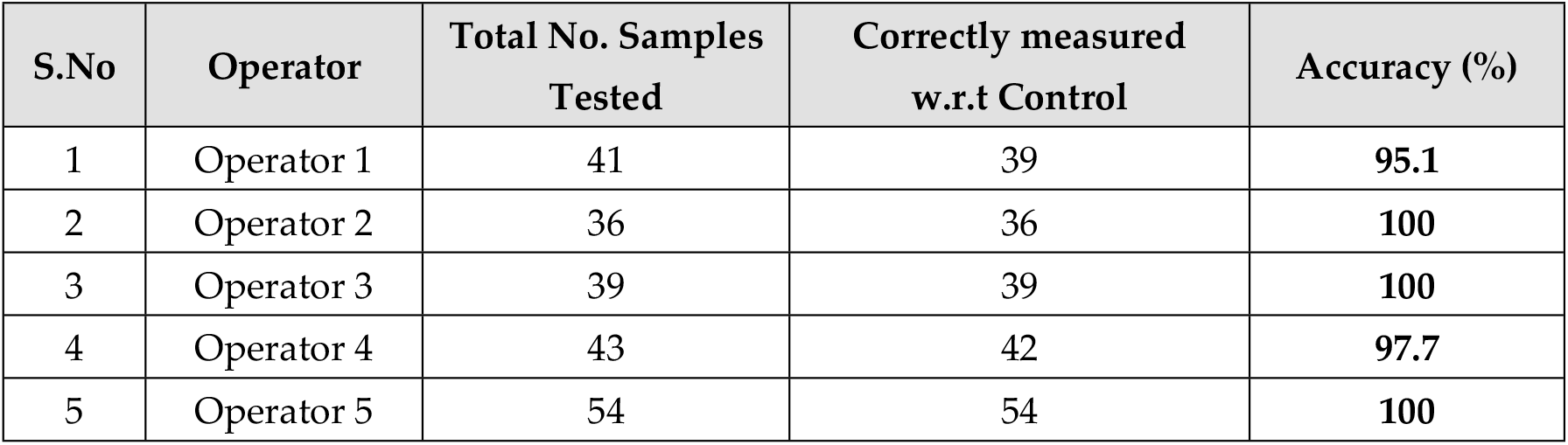
Test results for each untrained operator.

It is also observed that there is no reader instrument variability while interpreting the results when the samples are tested. All the RT-PCR comparative results correlate with the Kaya17 results when 5 different reader instruments were used for testing by 5 different untrained operators.

Figure 4 shows the inverse linear relationship between the observed Kaya17 RFUs versus the mean values of the N and ORF1ab Gene cycle thresholds (n=183, Pearson Correlation= -0.92). The figure shows that all of the negative samples by PCR were also Negative in Kaya17 nCoVega test, except for two samples (Sample Number 31and 117), all positive samples by PCR were also positive by Kaya17 nCoVega test, except for one sample (Sample Number 10).

**Figure 4:**
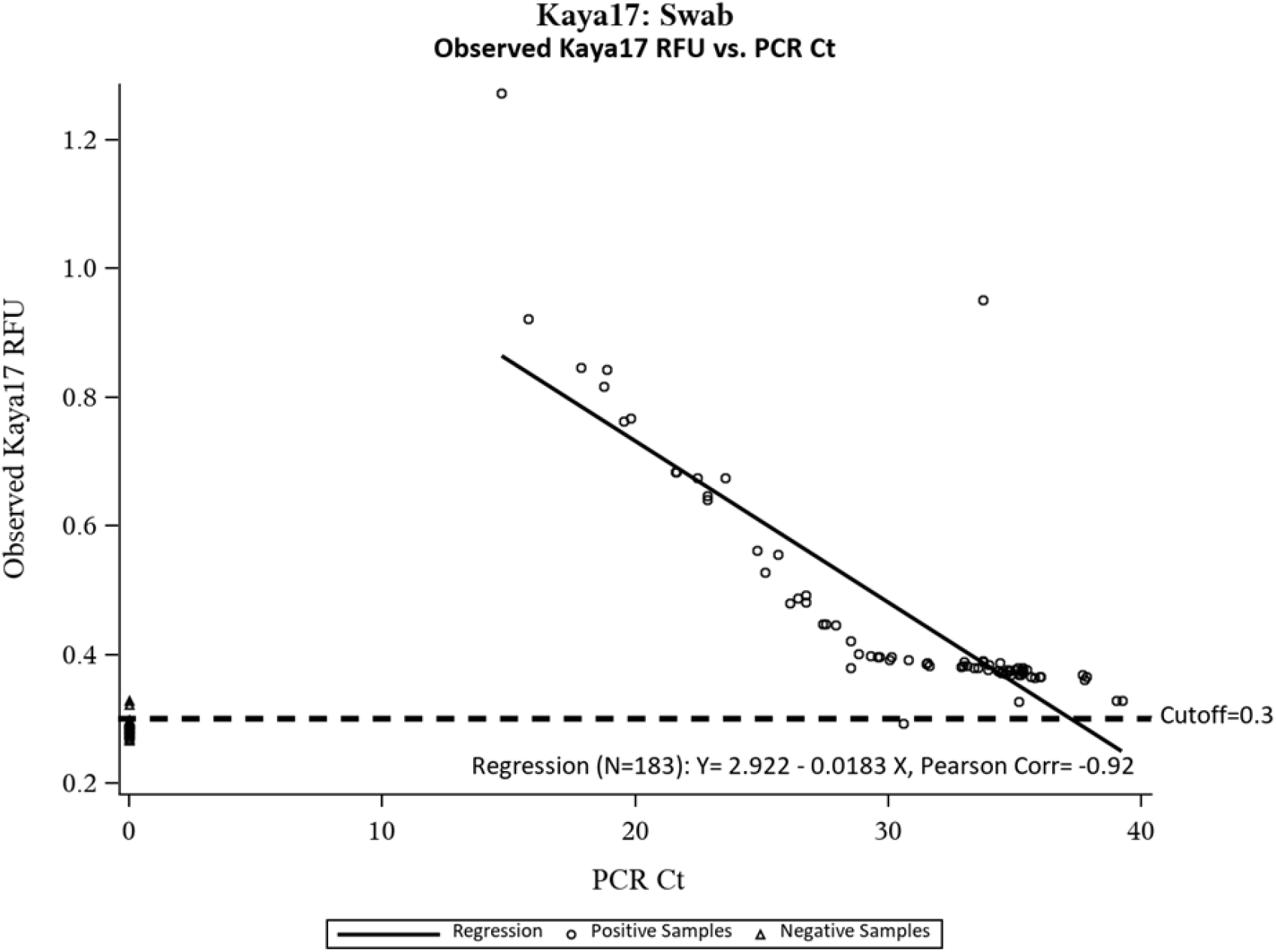
Inverse Relationship between Kaya17 RFUs vs Ct values from RT-PCR

## Discussion

The Kaya17 Vega-200 system and nCoVega antigen test is a simple-to-use, rapid assay that is also highly sensitive and specific. The test can detect very low levels of SARS-CoV-2 as seen in the high correlation with RT-PCR across all ranges of viral loads, all the way down to Ct counts of 38. The test also confirmed its capability to detect infected people as early as Day 1 from the start of symptoms. The reader instrument is light, portable and runs off USB-power from its connected laptop, so it does not need external power source. The system can also process 30-50 samples in one hour and can be run by untrained operators, making it a very useful test to apply at point of care sites for accurate detection of infected people very early before they spread the virus to others.The low LOD of 20 TCID_50_/ml validates a sensitivity that is simmilar to RT-PCR (7) and several orders of magnitiude more accurate/sensitive than most other antigen tests (13). A test like this with PCR-like sensitivity and accuracy has significant implications in surveillance and population control, especially given its use of small volume of saliva sample which is very easy and non-invasive for all ages of subjects. The software, instrument and disposables including cartridge, provide an end-to-end solution that can be deployed in mobile units, at schools, offices and events and also in remote areas, given the instrument’s small footprint and portability. Easily scalable manufacturing processes allow this test to meet any surge in demand for COVID-19 testing, as we open our schools, businesses and global travel and as new variants emerge. In the future, this test system can be productized for self-testing purposes. The nCoVega test can address the unmet need for rapid, accurate and inexpensive COVID-19 testing and it is suitable for broader public dissemination.

## Data Availability

All relevant data are within the manuscript

## Author Contributions

SD and VM conceived the study and cross-platform validation experiments at ASU. SS led the design and development of the Vega-200 system. SD, CBT, TC, AM, DM developed and optimized the nCoVega SARS-CoV-2 assay. DM and SD ran the analytical study. SD, CBT ran the clinical validation study at ASU. JC, JM and CBT analyzed the data. TC, SM and SD drafted the manuscript. CBT revised the manuscript. All authors read and approved the final manuscript.

## Institutional Review Board Statement

The study was conducted according to the guidelines of the Declaration of Helsinki, and approved by the Institutional Review Board (or Ethics Committee) of Western IRB (IRB protocol #20204097; Kaya17 protocol #KAYA-PROTOPOCT-007 Rev002) on 01/06/2021.

## Supplementary Materials

